# Machine Learning Based Detection of Chronic Kidney Disease using Full Blood Count from a Ghanaian Hospital Population with an Integrated Web Application

**DOI:** 10.64898/2026.10.27.26366677

**Authors:** Bismark Yao Akwetey, Eli Y. Lawluvi, Thomasia T. Equagoo, Sesinam Kumako, Esther A. Tsinyo, Rhodaline T. Kumi, Isaac R. Mensah, Franklin Amedume, Emmanuel Avuetor, Catherine E. Adikah, Winnefreda M. Mensah, Jeffrey Dwamena-Nkasah Botuo

## Abstract

Chronic kidney disease (CKD) is usually silent until advanced, and the tests confirming it are frequently unaffordable in sub-Saharan Africa, whereas the full blood count (FBC) is inexpensive and widely performed. We assessed whether machine learning applied to the FBC can detect CKD, and how far its reported performance depends on where synthetic minority oversampling (SMOTE) is applied. Records of 155 patients at the Ho Technical University Hospital, Ghana, 43 (27.7%) with CKD, yielded ten features: age, cardiovascular disease and eight raw FBC parameters. Eleven algorithms, six resampling strategies and thirteen ensembles were compared under repeated stratified cross-validation. With SMOTE applied before the train-test split, the protocol under which the deployed models were fitted, Random Forest and Extra Trees both reached an accuracy of 83.9%, with sensitivities of 0.893 and 0.857 and areas under the curve (AUC) of 0.925 and 0.939; repeated cross-validation gave an accuracy of 0.809 ± 0.051 and an AUC of 0.897. With resampling confined to the training folds, the same features gave a held-out accuracy of 61.5% and an AUC of 0.604 in 39 unseen patients. Age was the only variable separating the groups after correction for multiple testing (median 64 versus 47 years, adjusted p = 0.0026). The models are deployed as NephroScan GH (https://nephroscangh.com), which explains every prediction. The FBC carries a real but moderate signal for CKD, and the 22.4 point gap between the two protocols quantifies how far a single preprocessing decision can move a reported accuracy.

## Introduction

Chronic kidney disease affected an estimated 697.5 million people worldwide in 2017, a global prevalence of 9.1%, and caused 1.2 million deaths in that year; its all-age prevalence rose by 29.3% between 1990 and 2017 [1]. The burden falls unevenly. A systematic review of 90 studies from 96 sites in sub-Saharan Africa estimated a pooled CKD prevalence of 13.9% (95% CI 12.2–15.7) from the medium- and high-quality studies it identified [2]. In Ghana, a cross-sectional study of a high-risk population in the south-west reported CKD in 30% of those screened, rising to 74% among patients with both diabetes and hypertension [3]. The first annual report of the Ghana Renal Registry recorded 201 patients starting kidney replacement therapy in 2017, an incidence of 6.9 per million population, with hypertensive kidney disease the commonest primary cause at 39.9% and a median age at presentation of 45.5 years, two decades younger than in most high-income registries [4].

The diagnosis of CKD is not difficult in principle. The Kidney Disease: Improving Global Outcomes (KDIGO) 2024 guideline defines it as abnormalities of kidney structure or function present for more than three months, operationalised mainly through an eGFR below 60 mL/min/1.73 m² or a uACR of 30 mg/g or more [5]. The difficulty is access. An International Society of Nephrology survey of regional expert nephrologists drawn from all ten of the society’s regions found serum creatinine reported as available by every respondent and urine albumin by 96%, but with wide variation in what the tests cost and with reimbursement values that did not differ significantly between World Bank income groups [6]. Two things follow. The first is that an absolute price which is trivial in a high-income health system is not trivial in a Ghanaian one, where these tests are commonly paid for out of pocket. The second is that the respondents were expert nephrologists, that is, the best-resourced tier of the system; availability at a national referral centre says little about availability at a district hospital. In practice, a patient attending such a hospital may leave with a full blood count and without any measurement of kidney function at all, because the FBC is cheap, is performed on an automated analyser the hospital already owns, and is ordered almost reflexively for fever, fatigue, and antenatal care.

There are physiological reasons to expect the FBC to carry information about kidney function. Anaemia is a well-described consequence of declining erythropoietin production, with prevalence in the United States rising from 8.4% at CKD stage 1 to 53.4% at stage 5 [7]. Uraemia alters platelet size and function, and systemic inflammation, quantified in the FBC as the neutrophil-to-lymphocyte ratio, has been associated independently with progression to end-stage renal disease in a cohort of 938 patients with stages 1 to 4 CKD [8]. What is not established is whether these individual associations, taken together and in a population in which most detected disease is early, amount to a discriminative signal strong enough to be useful as a screening prompt.

Machine learning has been applied extensively to CKD. A rapid systematic review that screened 648 papers and included 68 found that the models performed well as reported by their authors, but that the metrics were too inconsistent between papers for direct comparison, that generalisability to diverse populations was rarely considered, and that only six of the 68 had been evaluated in a clinical context [9]. One widely cited study of the University of California, Irvine (UCI) CKD dataset reported 99.75% accuracy for a random forest and 99.83% for an integrated model [10]. Such figures are difficult to reconcile with the clinical reality of a disease that is routinely missed. Part of the explanation is the composition of the benchmark datasets, which commonly include serum creatinine, blood urea, and specific gravity, that is, the very measurements that define the outcome. Part of it is methodological: when synthetic minority oversampling is applied before, rather than after, the train–test split, synthetic points derived from test observations enter the training set and the resulting estimates are systematically optimistic, an effect that has been demonstrated directly for a medical prediction task [11]. Applied work has nevertheless begun to move towards deployment, with web-based CKD prediction tools [12] and explainable models that expose which inputs drive a prediction [13], and earlier work from our group has shown that a clinician-facing web platform can be built and operated in a Ghanaian hospital setting on low-cost infrastructure [14], consistent with the direction set out in the World Health Organization’s global strategy on digital health [15].

This study was designed around the test that patients in Ghana actually have in hand. We aimed to (1) establish whether machine learning applied to the FBC, age, and a short cardiovascular history can separate patients with CKD from patients without CKD in a Ghanaian hospital population, (2) quantify how much the reported performance of such a model depends on where synthetic minority oversampling is applied in the analysis pipeline, and (3) develop, explain, and deploy a web application that returns the resulting risk estimate to a non-specialist user together with the referral pathway that the estimate implies.

## Literature and Related Works

### Burden and Detection of CKD in Sub-Saharan Africa

CKD has been described as a public health priority for sub-Saharan Africa for more than a decade [16]. The epidemiological picture that has emerged since is one of a young patient population, a predominance of hypertensive and glomerular disease rather than diabetic nephropathy, and very limited access to kidney replacement therapy [2, 4]. Ghana’s registry data illustrate the consequence: 687 prevalent patients on kidney replacement therapy in 2017, a rate of 23.6 per million population, of whom 96.2% were receiving haemodialysis [4]. Where dialysis is paid for out of pocket, late detection is not merely a clinical problem but a financial one, and the case for inexpensive detection earlier in the disease course is correspondingly stronger.

### Haematological Changes in CKD

The haematological consequences of CKD are well characterised individually. Erythropoietin deficiency and shortened red cell survival produce a normocytic anaemia whose prevalence tracks the stage of disease [7]. Platelet indices shift as uraemic toxins accumulate, and mean platelet volume and platelet distribution width have been studied repeatedly as markers of this shift. Inflammation is a consistent feature, and the neutrophil-to-lymphocyte ratio has been associated with progression to end-stage renal disease, although in the C-STRIDE cohort that association reached significance only in stage 4 disease [8]. Taken individually these markers are neither sensitive nor specific. The question that motivates a multivariable approach is whether a model that reads all of them at once, along with their ratios and the patient’s age, recovers more signal than any one of them does alone.

### Machine Learning for CKD Detection

Reported performance in this literature is consistently high. The review of 68 studies found that models for CKD prognosis, diagnosis, and treatment management performed well as their authors reported them, while noting that the reported metrics were not comparable between studies, that generalisability was seldom examined, and that only six of the 68 studies were set in a clinical context [9]. Qin et al., working with the 400-record UCI dataset, reported 99.75% accuracy for a random forest after k-nearest-neighbour imputation and 99.83% for a perceptron-integrated model [10]. Ghosh and Khandoker, using 491 patient records and five algorithms, found XGBoost best and identified creatinine, glycated haemoglobin, and age as the most influential inputs [13]. Halder et al. went further towards practice, packaging a CKD classifier behind a web front end so that the model could be exercised by a user rather than only by a reviewer [12].

Two features of this body of work bear directly on the present study. The first is input composition: models that include serum creatinine, blood urea, or urine protein are not solving the problem faced by a hospital that cannot measure them. The second is protocol. Vandewiele et al., working on preterm-birth prediction from electrohysterogram data, attributed the near-perfect results repeatedly reported on that benchmark to oversampling applied before partitioning, demonstrated the bias on two artificial datasets, reproduced the flawed results, and then re-evaluated the same methods with the oversampler applied after partitioning to obtain a realistic estimate [11]. The mechanism belongs to the broader family of leakage effects catalogued by Kaufman et al., in which information from the evaluation set reaches the learner through the preparation of the data rather than through the features themselves [17]. SMOTE itself was introduced as a way of addressing class imbalance by interpolating between minority neighbours [18]; the interpolation is precisely what makes its placement in the pipeline consequential, because a synthetic point generated from a test observation is a partial copy of it.

### Reporting Standards and Sample Size

The TRIPOD+AI statement sets out what a prediction-model study should report, including the handling of class imbalance, the separation of development from evaluation data, and the presentation of calibration alongside discrimination [19]. Riley et al. provide the complementary quantitative constraint: the minimum sample size needed to develop a prediction model with acceptable shrinkage and precision, a calculation which for a binary outcome depends on the number of candidate predictors, the outcome prevalence, and the anticipated model performance [20]. Both documents are relevant to a single-centre study of 155 patients.

## Materials and Methods

### Study Design, Setting, and Ethical Approval

This was a retrospective, single-centre, cross-sectional study of patients attending The Ho Technical University Hospital, Ho, Volta Region, Ghana. Ethical clearance was obtained from the Directorate of Research and Innovation, Ho Technical University, under approval number HTU/DRI/EC2026-071. Records were de-identified at extraction; no name, hospital number, address, or date of birth was transferred to the analysis dataset, and the authors who performed the analysis had no access to identifiers.

### Participants and Variables

Records of 156 patients were extracted. For each patient the dataset held 28 fields: five binary descriptors (sex, diabetes, anaemia, cardiovascular disease, and hypertension), four clinical measurements (age, weight, systolic and diastolic blood pressure), 18 full blood count parameters, and the recorded CKD level. The FBC parameters were red blood cell count (RBC), haemoglobin (HGB), haematocrit (HCT), mean corpuscular volume (MCV), mean corpuscular haemoglobin (MCH), mean corpuscular haemoglobin concentration (MCHC), red cell distribution width expressed as a coefficient of variation (RDW_cv) and as a standard deviation (RDW_sd), mixed cell count (MXD), white blood cell count (WBC), neutrophils (NEUT), lymphocytes (LYM), platelet count (PLT), mean platelet volume (MPV), plateletcrit (PCT), platelet large cell ratio (PLCR), platelet large cell count (PLCC), and platelet distribution width (PDW). The CKD level was recorded on the hospital’s six-point scale as NORMAL, MILD CKD, MODERATE CKDA, MODERATE CKDB, SEVERE CKD, or END STAGE.

### Data Cleaning

Four numeric fields had been entered with a decimal comma rather than a decimal point and were parsed as text (MCH in one row, MXD in one row, NEUT in one row, and MPV in one row); these were converted. The file contained no missing values and no exactly duplicated rows. A search for near-duplicates, defined as pairs of records agreeing on at least 14 FBC values, returned one pair (rows 118 and 124), which differed only in weight, blood pressure, RDW_sd, NEUT, PCT, and PDW while agreeing on 14 other haematological values; the second record was dropped as a probable duplicate entry, leaving 155 patients.

We then applied three internal-consistency rules that the FBC must satisfy by construction: MCV must equal HCT/RBC × 10, MCH must equal HGB/RBC × 10, and MCHC must equal HGB/HCT × 100; the white cell differential (NEUT + LYM + MXD) must sum to the WBC within tolerance; and the platelet indices must satisfy PCT = PLT × MPV / 10 000 and PLCC = PLT × PLCR / 100. A fourth rule required the diastolic pressure to be lower than the systolic pressure; no record violated it. Where a value violated a rule and could be re-derived unambiguously from the others within a physiologically plausible window, it was replaced by the derived value. Where the faulty value was ambiguous, or where it was physiologically impossible, it was set to missing and left to be imputed inside the modelling pipeline. This produced 17 value-level corrections in 14 rows: two MCV values re-derived, three MCHC or MCH values corrected or voided, one RDW_sd voided, six differential values re-derived and two voided, one MPV re-derived, one PLT re-derived, and two platelet indices voided. Seven missing values across seven columns resulted and were imputed by the column median within each cross-validation fold, never across folds.

### Outcome Definition

The six-point CKD level was collapsed into a binary outcome: CKD of any stage (MILD, MODERATE A, MODERATE B, SEVERE, or END STAGE) versus no CKD. This choice reflects the intended use. A screening aid is useful if it tells a patient to obtain an eGFR and a uACR; it does not need to stage the disease, and with only three end-stage and two severe cases in the cohort, a multiclass model could not have been fitted responsibly.

### Feature Engineering

Ten derived variables were computed from the raw fields, each with an established haematological or clinical rationale: the neutrophil-to-lymphocyte ratio (NLR), platelet-to-lymphocyte ratio (PLR), systemic immune-inflammation index (SII = PLT × NEUT / LYM), monocyte-to-lymphocyte ratio (MLR, computed from MXD), red cell distribution width-to-platelet ratio (RPR), haemoglobin-to-red cell distribution width ratio (HRR), MPV-to-platelet ratio, the Mentzer index (MCV/RBC), pulse pressure, and mean arterial pressure. With the 27 raw predictors this gave 37 candidate features.

### Exploratory Data Analysis

Distributions were summarised by count, mean, standard deviation, minimum, quartiles, and maximum. Because most FBC parameters were skewed and the CKD group was small, the two groups were compared with the Mann-Whitney U test rather than Student’s t test, and effect sizes were reported as the rank-biserial correlation r. All 32 numeric comparisons were corrected for multiple testing by the Benjamini-Hochberg procedure, controlling the false discovery rate at 5% [21]. Categorical comorbidities were compared as prevalence by class.

### Data Partitioning and Cross-Validation

The 155 patients were split once into a training set of 116 and a held-out test set of 39, stratified on the outcome, giving 84 and 28 patients without CKD and 32 and 11 patients with CKD, respectively. The test set was touched only once, at the end, for the final evaluation. All model selection, resampling, tuning, and threshold choice were performed inside 5-fold stratified cross-validation repeated five times on the training set, that is, 25 fits per configuration.

### Two Resampling Protocols

The class imbalance (27.7% CKD) was addressed by synthetic minority oversampling [18]. We deliberately evaluated two protocols, because the comparison between them is one of the study’s results.

#### SMOTE Inside Training Only (Conservative Protocol)

Imputation, scaling, and oversampling were placed inside a pipeline that was refitted from scratch on each training fold, so that no synthetic point was ever derived from an observation used to score that fold. Six samplers were compared: no resampling, SMOTE, Borderline-SMOTE, ADASYN [22], SMOTE-Tomek [23], and SMOTE-ENN.

#### SMOTE Before the Split (Augmented Protocol)

SMOTE was applied once to all 155 patients, producing 224 rows (112 without CKD and 112 with CKD, that is, 69 synthetic CKD rows), and the augmented set was then split 75:25 into 168 training and 56 test rows. This is the protocol under which the models now in production were fitted. Because the oversampler interpolates between a minority observation and its neighbours, synthetic rows in the training partition can be partial copies of patients in the test partition; the evaluation is therefore optimistic by construction, and we report both the inflated figure and its decomposition into synthetic and real test rows.

### Feature Selection

Within the training set, pairs of features with an absolute Spearman correlation above 0.9 were identified and the member of each pair with the lower mutual information with the outcome was dropped, removing HGB, HRR, MAP, MPV_PLT, PCT, PLCR, RPR, and SII and leaving 29 candidates. The survivors were then ranked by three independent criteria, mutual information, the ANOVA F statistic, and out-of-fold permutation importance from a random forest averaged over a 5-fold, two-repeat cross-validation with 10 permutations per fold, and ordered by mean rank. The cross-validated ROC AUC of a SMOTE-plus-random-forest pipeline was then computed for the top k features for every k from 2 to 29, and k was chosen by the one-standard-error rule applied to a three-point moving average.

A domain constraint was imposed on the result. Because the application is intended to run on a full blood count, we required the final feature set to contain at least eight raw FBC parameters, adding the next-highest-ranked FBC parameters until that floor was met. The final set of ten features was therefore age, cardiovascular disease, and the eight FBC parameters WBC, MPV, PLCC, MCHC, MXD, PDW, PLT, and NEUT.

### Algorithm Screening, Tuning, and Ensembling

Eleven algorithms were screened under the conservative protocol with default settings: logistic regression, k-nearest neighbours, a support vector machine with a radial basis function kernel, a decision tree, random forest [24], extra trees [25], gradient boosting, histogram-based gradient boosting, AdaBoost, Gaussian naive Bayes, and a multilayer perceptron. The three strongest by cross-validated ROC AUC were carried forward. For each, the best of the six samplers was selected and hyperparameters were tuned by randomised search over 25 candidate configurations with the same repeated cross-validation. The tuned models were then combined by soft voting and by stacking with a logistic-regression meta-learner.

A second family of ensembles was evaluated independently: bagged decision trees, bagged k-nearest neighbours, bagged support vector machines, bagged logistic regression, bagged gradient boosting, balanced bagging with SMOTE applied within each bag, balanced random forest [26], EasyEnsemble [27], and RUSBoost [28], followed by soft voting and stacking over the six strongest members.

### Decision Threshold

Because the cohort is imbalanced and a missed case of CKD costs more than an unnecessary laboratory referral, the default 0.5 threshold was not retained. Out-of-fold predicted probabilities from the training set were swept from 0.05 to 0.95 and the threshold maximising balanced accuracy was selected. No test-set information entered this choice.

### Evaluation Metrics

Accuracy, balanced accuracy, sensitivity, specificity, precision, F1, ROC AUC, average precision, Matthews correlation coefficient (MCC), and Cohen’s kappa were computed. MCC is reported throughout because it is the metric least distorted by class imbalance [28]. Ninety-five percent confidence intervals for the held-out metrics were obtained by bootstrap resampling of the test set with 2000 replicates. The majority-class baseline, always predicting no CKD, is reported alongside accuracy so that accuracy cannot be read in isolation.\

### Model Explanation

For a forest, the predicted probability equals the probability at the root plus the sum of the probability changes along each decision path, so every split can be attributed exactly to the feature it tested. We used this decomposition, rather than an approximation, to express each prediction as a bias term plus one signed contribution per input [29], and verified the identity numerically: the sum of the bias and the ten contributions reproduced the predicted probability to within floating-point error. Permutation importance on the held-out set was computed separately as a global summary.

### Web Application

NephroScan GH was implemented as a FastAPI service with an unbundled HTML, CSS, and JavaScript front end totalling 98.4 KB, served with gzip compression and no client-side framework. The trained pipelines, 1.75 MB in total, are loaded once at process start. Nine endpoints are exposed: a health check, a model-description endpoint, the prediction endpoint, a town index, a renal-centre directory, a geocoding helper, a care-finder, a laboratory finder, and the single-page front end. The prediction endpoint validates the ten inputs with Pydantic against clinically plausible bounds that are deliberately wider than the training ranges; a value outside the training range is flagged in the response rather than rejected, so that the user is told the model is extrapolating instead of being refused an answer.

The clinical interface was built around three decisions. First, the two deployed forests are scored on every request and the user is shown their average, not a choice of models, under the plain-language names Sentinel (Extra Trees) and Guardian (Balanced Random Forest); the technical identity of each is given in the interface, but the reported number is a single score. Second, the decision-path contributions are rendered as signed horizontal bars, scaled so that the largest contribution in a given result fills the available width, with an explicit legend stating that a longer bar means a larger effect. Third, the score is banded into low (below 0.35), moderate (0.35 to 0.50), and high (0.50 and above) risk, the upper cut-off being the decision threshold recorded in the deployment metadata, and every band carries the same instruction to obtain a serum creatinine with eGFR and a urine albumin-to-creatinine ratio. A reference-range check against typical adult values is displayed for each FBC input, with a note that laboratory ranges differ; this check never alters the prediction. A directory of 47 towns, 10 renal and referral centres, and 27 laboratories supports the referral step, with directions resolved through a live map search by facility name rather than by stored coordinates.

### Deployment

The application is deployed from a Git repository to a managed container platform, built with `pip install -r requirements.txt` and started with `uvicorn app.main:app --host 0.0.0.0 - -port $PORT` on Python 3.11.11, with the health endpoint used as the platform’s readiness probe and a custom domain, nephroscangh.com, mapped through DNS. Deployment is automatic on push to the main branch.

### Software

Analyses were performed in Python 3.11.15 with scikit-learn 1.8.0 [30], imbalanced-learn 0.15.dev0 [31], pandas 3.0.2, numpy 2.4.4, and matplotlib. The scikit-learn version is pinned in the deployment requirements because it is the version under which the production pipelines were serialised. The complete analysis is provided as an executable Jupyter notebook (see Data Availability), and every figure and numerical value reported below is produced by that notebook.

## Results

### Cohort Characteristics

After cleaning, 155 patients remained. One hundred and twelve (72.3%) had no CKD and 43 (27.7%) had CKD of any stage; within the CKD group, 23 were mild, 10 moderate A, five moderate B, two severe, and three end stage. The majority-class baseline, always predicting no CKD, therefore corresponds to an accuracy of 72.3% in the full cohort and 71.8% in the held-out test set, and no model should be credited for accuracy below those values.

Mean age was 51.9 ± 17.1 years (range 7 to 97), mean weight 77.0 ± 16.7 kg, mean systolic blood pressure 145.7 ± 27.0 mmHg, and mean diastolic blood pressure 92.6 ± 19.7 mmHg. Thirty-three point five percent of the cohort were male. Hypertension was recorded in 66.5%, anaemia in 34.8%, cardiovascular disease in 27.7%, and diabetes in 14.2%. The high prevalence of hypertension is consistent with the registry finding that hypertensive kidney disease is the commonest primary diagnosis in Ghana [4], and the low prevalence of diabetes distinguishes this cohort from the cohorts on which most published CKD models were developed. Descriptive statistics for all 23 numeric variables and the ten engineered ratios are given in Table 1.

**Table 1:** Descriptive statistics of the 155 patients after cleaning. Counts below 155 reflect values set to missing by the consistency checks described in Section 3.3 and imputed within the modelling pipeline.

| Variable | Unit | n | Mean | SD | Min | Q1 | Median | Q3 | Max |
|---|---|---|---|---|---|---|---|---|---|
| Age | years | 155 | 51.86 | 17.05 | 7.00 | 40.00 | 50.00 | 64.00 | 97.00 |
| Weight | kg | 155 | 77.02 | 16.68 | 32.00 | 67.00 | 75.00 | 88.00 | 126.00 |
| Systolic BP | mmHg | 155 | 145.68 | 26.97 | 103.00 | 126.50 | 140.00 | 158.50 | 258.00 |
| Diastolic BP | mmHg | 155 | 92.57 | 19.67 | 61.00 | 78.50 | 88.00 | 99.50 | 180.00 |
| RBC | ×10<sup>12</sup>/L | 155 | 4.43 | 0.72 | 1.95 | 4.11 | 4.50 | 4.88 | 6.03 |
| HGB | g/dL | 155 | 12.27 | 2.15 | 5.20 | 11.40 | 12.50 | 13.80 | 16.10 |
| HCT | % | 155 | 36.70 | 5.84 | 17.40 | 34.85 | 37.30 | 40.40 | 49.50 |
| MCV | fL | 155 | 82.81 | 7.30 | 59.56 | 78.90 | 83.60 | 87.65 | 108.80 |
| MCH | pg | 154 | 27.66 | 3.08 | 17.50 | 26.00 | 28.00 | 29.68 | 36.70 |
| MCHC | g/dL | 154 | 33.26 | 1.68 | 28.10 | 32.20 | 33.35 | 34.40 | 37.80 |
| RDW_cv | % | 155 | 13.69 | 1.50 | 11.20 | 12.70 | 13.40 | 14.25 | 19.70 |
| RDW_sd | fL | 154 | 44.48 | 3.93 | 30.60 | 41.90 | 44.30 | 46.18 | 59.10 |
| MXD | ×10<sup>9</sup>/L | 154 | 0.84 | 0.36 | 0.23 | 0.62 | 0.76 | 0.98 | 3.13 |
| WBC | ×10<sup>9</sup>/L | 155 | 6.72 | 2.53 | 2.94 | 5.26 | 6.08 | 7.52 | 21.11 |
| NEUT | ×10<sup>9</sup>/L | 154 | 3.57 | 2.40 | 0.80 | 2.04 | 2.90 | 4.44 | 18.27 |
| LYM | ×10<sup>9</sup>/L | 155 | 2.32 | 0.86 | 0.43 | 1.78 | 2.35 | 2.84 | 4.86 |
| PLT | ×10<sup>9</sup>/L | 155 | 251.37 | 93.06 | 59.00 | 195.50 | 246.00 | 286.50 | 717.00 |
| MPV | fL | 155 | 10.00 | 0.90 | 8.20 | 9.40 | 9.90 | 10.60 | 13.50 |
| PCT | % | 155 | 0.25 | 0.08 | 0.06 | 0.20 | 0.24 | 0.28 | 0.60 |
| PLCR | % | 154 | 21.16 | 6.22 | 9.50 | 16.52 | 20.40 | 25.55 | 45.00 |
| PLCC | ×10<sup>9</sup>/L | 154 | 50.85 | 18.71 | 13.00 | 38.00 | 49.00 | 57.00 | 115.00 |
| PDW | fL | 155 | 13.57 | 2.10 | 9.50 | 12.05 | 13.30 | 14.80 | 19.70 |
| NLR | ratio | 154 | 2.03 | 2.16 | 0.32 | 0.78 | 1.19 | 2.42 | 14.50 |
| PLR | ratio | 155 | 127.59 | 81.42 | 12.14 | 81.54 | 106.67 | 141.27 | 642.59 |
| SII | index | 154 | 508.20 | 627.48 | 50.60 | 182.98 | 301.63 | 536.98 | 5336.00 |
| MLR | ratio | 154 | 0.44 | 0.35 | 0.11 | 0.24 | 0.33 | 0.49 | 2.19 |
| RPR | ratio | 155 | 6.26 | 3.01 | 1.77 | 4.65 | 5.48 | 6.96 | 25.59 |
| HRR | ratio | 155 | 0.91 | 0.20 | 0.33 | 0.82 | 0.93 | 1.05 | 1.37 |
| MPV/PLT | ratio | 155 | 4.64 | 2.34 | 1.16 | 3.35 | 3.93 | 5.31 | 18.81 |
| Mentzer index | ratio | 155 | 19.44 | 5.35 | 11.44 | 16.66 | 18.52 | 20.78 | 55.79 |
| Pulse pressure | mmHg | 155 | 53.11 | 14.90 | 25.00 | 43.00 | 51.00 | 61.00 | 119.00 |
| MAP | mmHg | 155 | 110.28 | 21.24 | 80.00 | 96.00 | 105.33 | 119.83 | 206.00 |

### Univariable Comparisons

Table 2 reports the ten variables with the largest effect sizes from the Mann–Whitney comparisons, and Figure 1 shows the distributions of twelve representative FBC parameters, FBC ratios, and clinical variables by CKD status. Age was the only variable to survive correction for multiple comparisons: the median age was 64 years in the CKD group and 47 years in the group without CKD (adjusted p = 0.0026, rank-biserial r = 0.4099). Among the FBC parameters, the largest uncorrected differences were in neutrophils (median 3.45 versus 2.64 × 10⁹/L, r = 0.2753, p = 0.0087), the neutrophil-to-lymphocyte ratio (1.42 versus 1.06, r = 0.2368, p = 0.0240), white blood cells (6.50 versus 5.97 × 10⁹/L, r = 0.2342, p = 0.0243), mean platelet volume (9.60 versus 10.00 fL, r = −0.2236, p = 0.0314), and large platelet count (44.0 versus 49.0 × 10⁹/L, r = −0.2122, p = 0.0430), none of which remained significant after adjustment (adjusted p = 0.139 to 0.230). Refer to Table2 and Fig1

**Figure 1:**
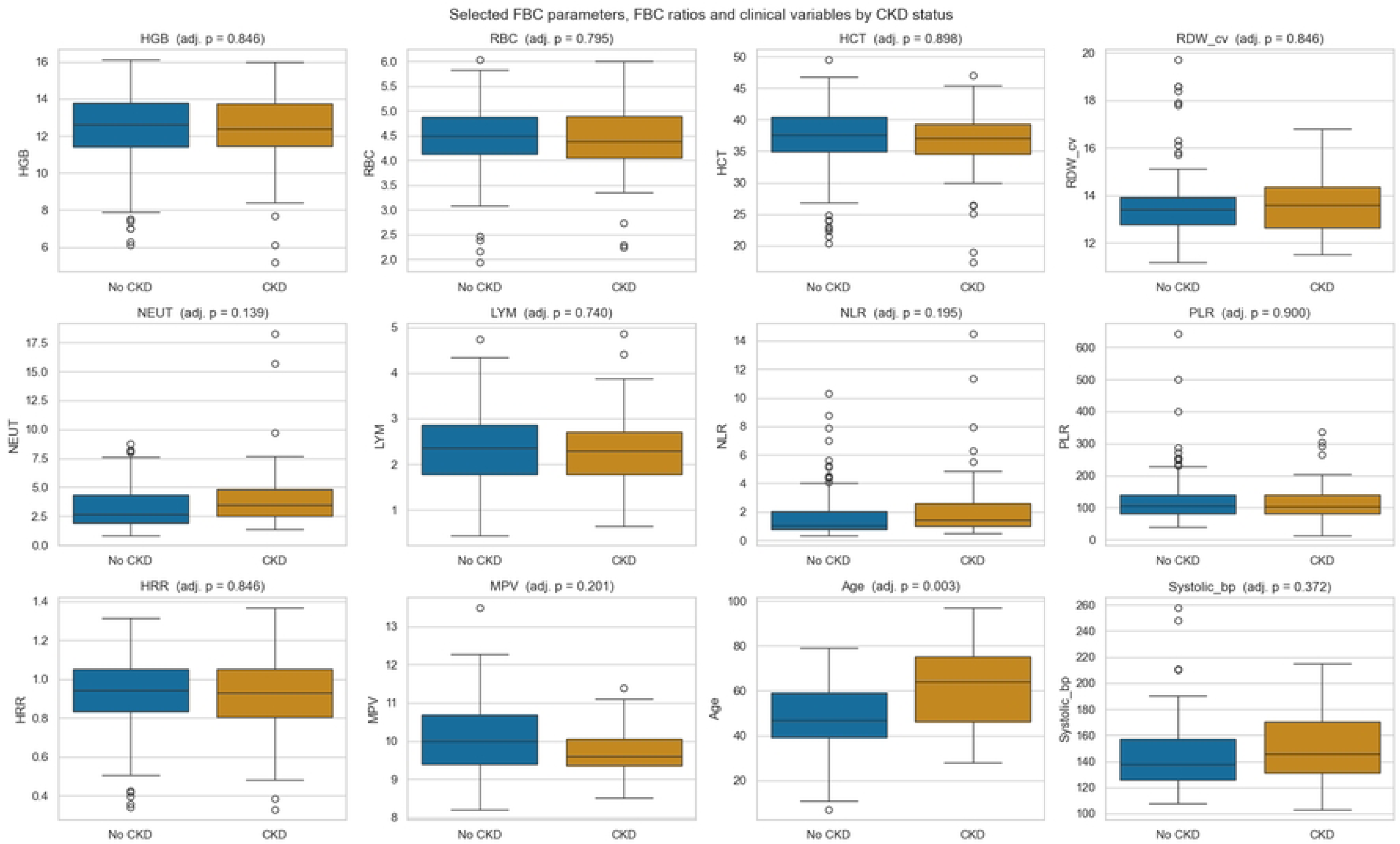
Distributions of selected full blood count parameters, engineered FBC ratios, and clinical variables by CKD status, with Benjamini–Hochberg adjusted p values. Age is the only panel in which the two distributions separate.

**Table 2:** Mann–Whitney U comparisons between patients with and without CKD, ordered by the absolute rank-biserial effect size. The ten strongest of 32 numeric comparisons are shown. P values were adjusted for multiple testing by the Benjamini–Hochberg procedure.

| **Variable** | **Group** | **Median, no CKD** | **Median, CKD** | **Effect size *r*** | ***p*** | **Adjusted *p*** |
|---|---|---|---|---|---|---|
| Age | Clinical | 47.00 | 64.00 | 0.4099 | 0.0001 | 0.0026 |
| NEUT | FBC | 2.64 | 3.45 | 0.2753 | 0.0087 | 0.1387 |
| NLR | FBC ratio | 1.063 | 1.424 | 0.2368 | 0.0240 | 0.1945 |
| WBC | FBC | 5.97 | 6.50 | 0.2342 | 0.0243 | 0.1945 |
| MPV | FBC | 10.00 | 9.60 | −0.2236 | 0.0314 | 0.2009 |
| PLCC | FBC | 49.00 | 44.00 | −0.2122 | 0.0430 | 0.2296 |
| Systolic BP | Clinical | 138.00 | 146.00 | 0.1736 | 0.0951 | 0.3720 |
| Pulse pressure | Clinical | 50.50 | 56.00 | 0.1730 | 0.0963 | 0.3720 |
| PLCR | FBC | 20.65 | 19.45 | −0.1703 | 0.1046 | 0.3720 |
| SII | FBC ratio | 278.70 | 366.97 | 0.1458 | 0.1647 | 0.4879 |

Two negative findings deserve emphasis. Haemoglobin did not differ between the groups (median 12.4 versus 12.6 g/dL, adjusted p = 0.846), and neither did haematocrit, red cell count, or the red cell distribution width. Anaemia, recorded as a clinical diagnosis, was present in 34.9% of the CKD group and 34.8% of the group without CKD. This is not inconsistent with the literature: anaemia in CKD becomes prevalent as filtration falls [7], and 33 of the 43 CKD cases in this cohort were mild or moderate. The corollary is that the red cell indices, the part of the FBC most often assumed to carry the renal signal, contributed almost nothing here, and the discriminative information came instead from the white cell and platelet compartments.

Comorbidity prevalences by class are given in Table 3. Cardiovascular disease was more than twice as common in the CKD group (46.5% versus 20.5%) and hypertension was more common (81.4% versus 60.7%), while diabetes was slightly less common (11.6% versus 15.2%). Refer to Table3

**Table 3:** Prevalence of recorded comorbidities and sex by CKD status (n = 155).

| **Group** | **Male (%)** | **Diabetes (%)** | **Anaemia (%)** | **Cardiovascular disease (%)** | **Hypertension (%)** |
|---|---|---|---|---|---|
| CKD (n = 43) | 37.2 | 11.6 | 34.9 | 46.5 | 81.4 |
| No CKD (n = 112) | 32.1 | 15.2 | 34.8 | 20.5 | 60.7 |

### Redundancy Pruning and Feature Ranking

Ten feature pairs exceeded the 0.9 correlation threshold, and eight features were dropped as redundant: HGB (with HCT, ρ = 0.945), HRR (with HGB, ρ = 0.908), MAP (with diastolic and systolic blood pressure, ρ = 0.955 and 0.926), MPV_PLT (with PLT, ρ = 0.957), PCT (with PLT, ρ = 0.951), PLCR (with MPV, ρ = 0.952), RPR (with PLT, ρ = 0.923), and SII (with NLR, ρ = 0.909). Twenty-nine candidates remained.

The consensus ranking placed age first (mean rank 1.33), cardiovascular disease second (4.33), and then WBC (4.67), MPV and PLCC (5.67 each), MCHC (8.33), MXD (8.67), PDW (9.00), and PLT (9.67). Figure 2 shows the cross-validated ROC AUC as a function of the number of top-ranked features retained. The curve rises steeply to k = 5, peaks around k = 9, and declines thereafter; the one-standard-error rule selected k = 7. Applying the eight-parameter FBC floor added PDW, PLT, and NEUT, giving the final ten features. The constraint improved rather than degraded performance: the cross-validated ROC AUC was 0.747 ± 0.092 for the seven data-selected features, 0.772 ± 0.074 for the final ten, and 0.688 ± 0.077 for all 29 candidates (Table 4 and Fig2).

**Figure 2:**
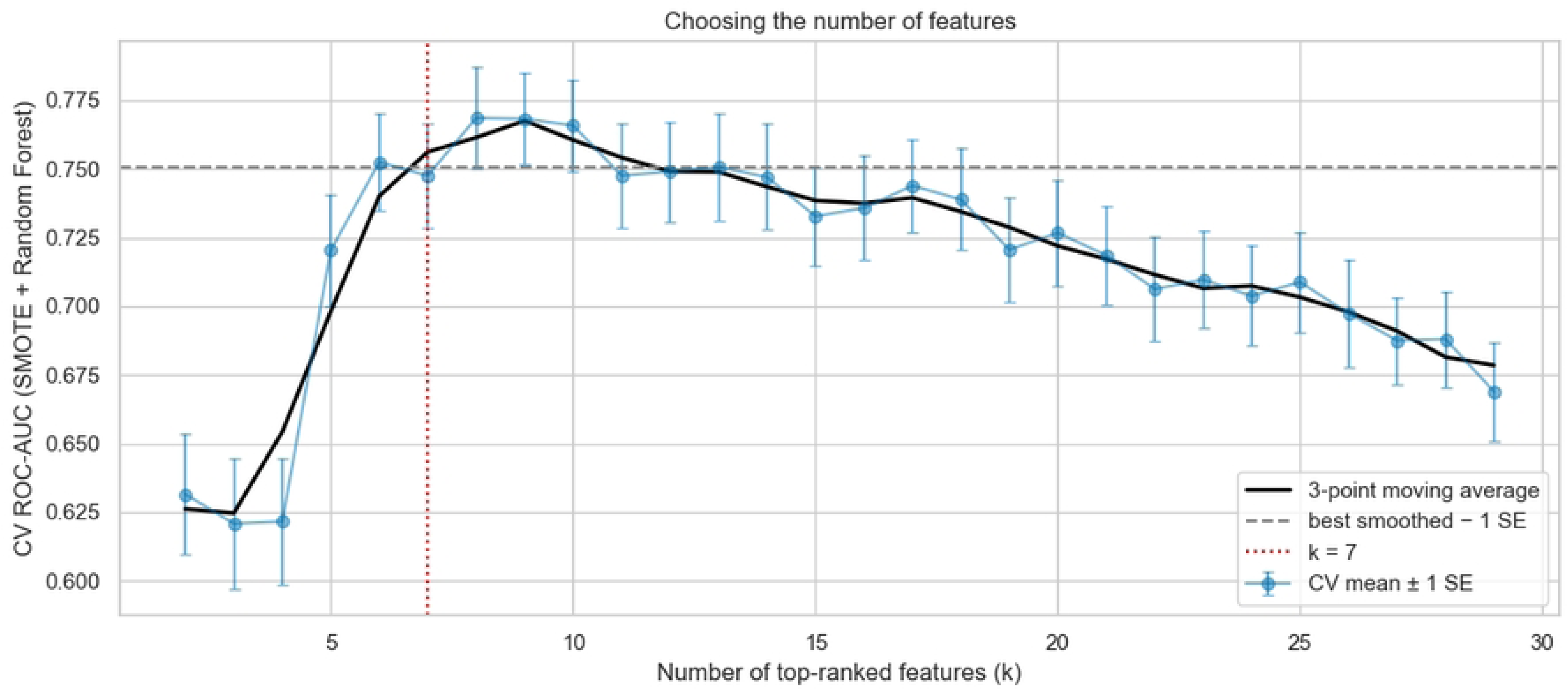
Cross-validated ROC AUC of a SMOTE-plus-random-forest pipeline as a function of the number of top-ranked features retained, with the one-standard-error rule selecting k = 7. Error bars are one standard error across the 25 fits.

**Table 4:** Cross-validated ROC AUC of three candidate feature sets, computed on the training set under 5-fold stratified cross-validation repeated five times.

| **Feature set** | **Number of features** | **CV ROC AUC** |
|---|---|---|
| Data-driven selection | 7 | 0.747 ± 0.092 |
| Final set with the FBC floor | 10 | 0.772 ± 0.074 |
| All candidate features | 29 | 0.688 ± 0.077 |

### Algorithm Screening

Table 5 and Figure 3 report the cross-validated performance of the eleven algorithms under the conservative protocol. Random forest was best by ROC AUC (0.772 ± 0.074), followed by histogram-based gradient boosting (0.771 ± 0.096) and gradient boosting (0.754 ± 0.064); the decision tree was worst (0.586 ± 0.095). The pattern across the table is informative in itself: the four forest and gradient-boosting methods achieved high specificity (0.814 to 0.863) and low sensitivity (0.396 to 0.564) at the default threshold, while k-nearest neighbours and naive Bayes inverted that trade-off, reaching sensitivities of 0.71 and 0.67 at specificities of 0.64 and 0.63. No configuration achieved both. (See Table5 and Fig3)

**Figure 3:**
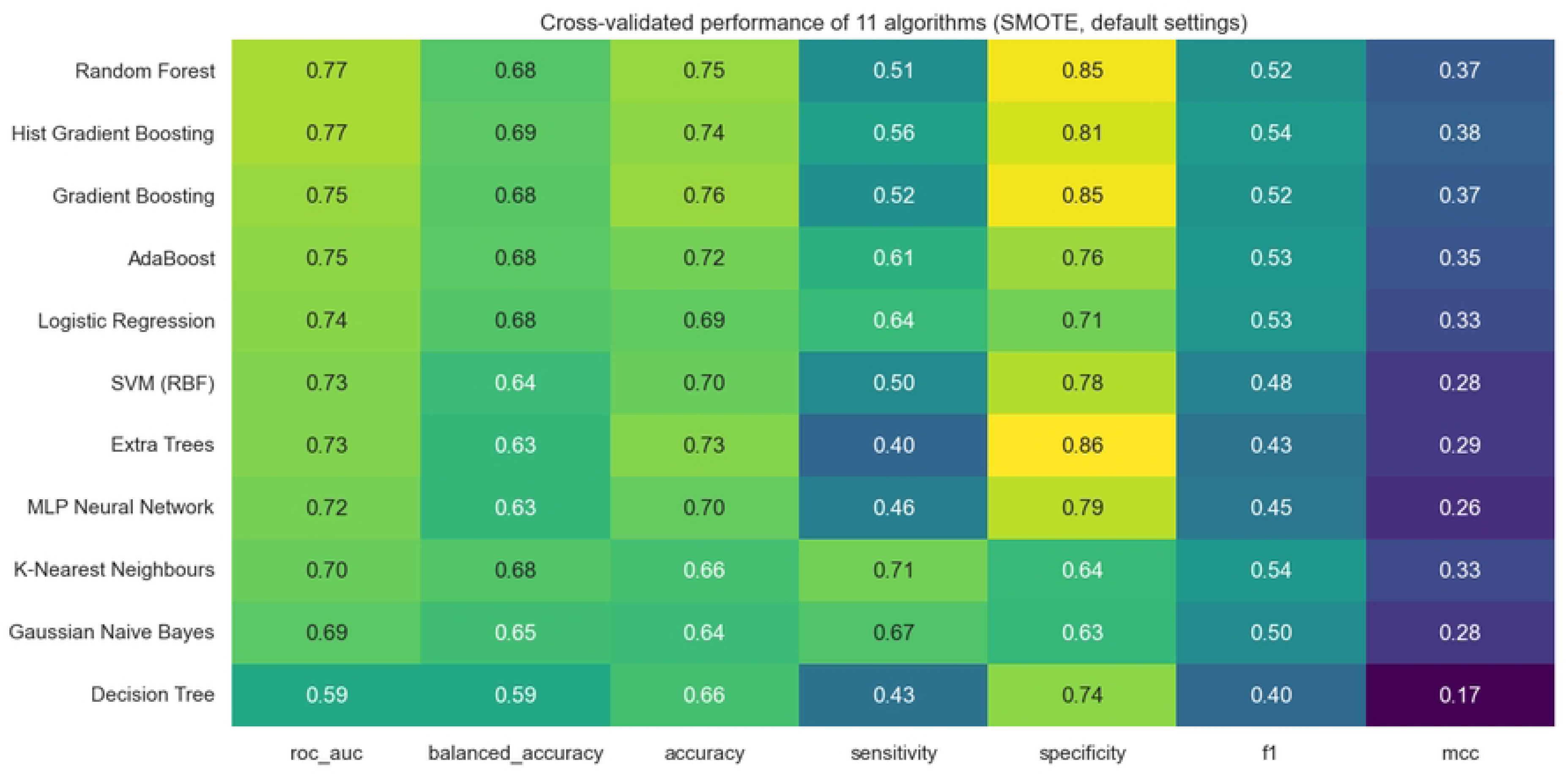
Cross-validated performance of the eleven screened algorithms under the conservative protocol, ordered by ROC AUC.

**Table 5:**
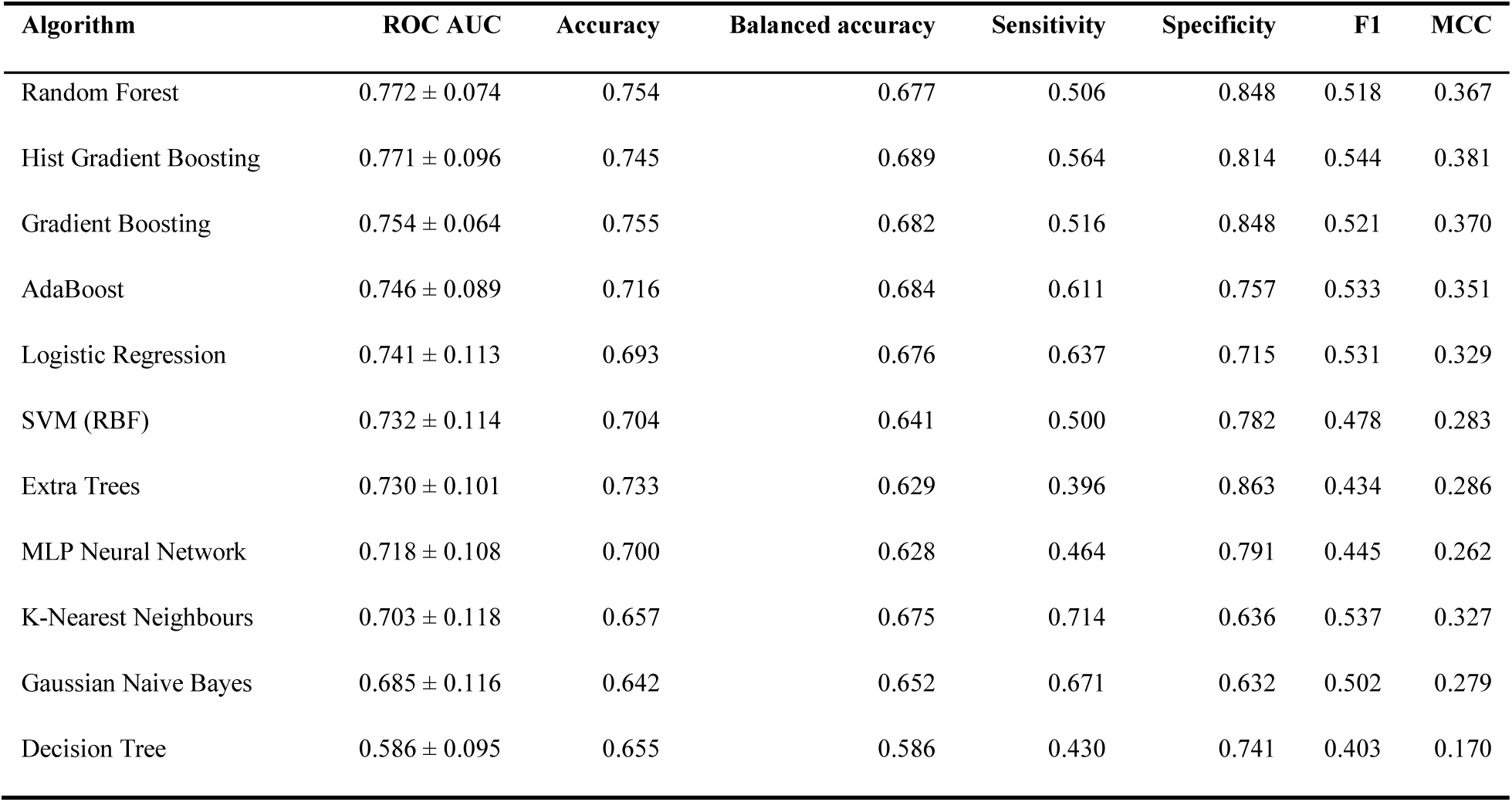
Cross-validated performance of eleven algorithms with default settings under the conservative protocol (SMOTE inside the training folds only), ordered by ROC AUC. All metrics are means over 25 fits at the default 0.5 threshold. The majority-class baseline gives an accuracy of 0.724, a balanced accuracy of 0.500, and a ROC AUC of 0.500.

### Resampling and Tuning

Figure 4 (Fig4) compares the six samplers across the three strongest algorithms. Differences were small and not consistent in direction: for random forest, SMOTE-Tomek gave the best ROC AUC (0.773) against 0.766 with no resampling at all; for histogram-based gradient boosting, ADASYN was best (0.772 against 0.763); for gradient boosting, SMOTE-Tomek was best (0.761 against 0.759). Resampling had a clearer effect on balanced accuracy, raising it from 0.596 with no resampling to 0.677 with SMOTE for random forest, and from 0.650 to 0.689 for histogram-based gradient boosting, which is the expected consequence of shifting the implicit operating point rather than of improved ranking.

**Figure 4:**
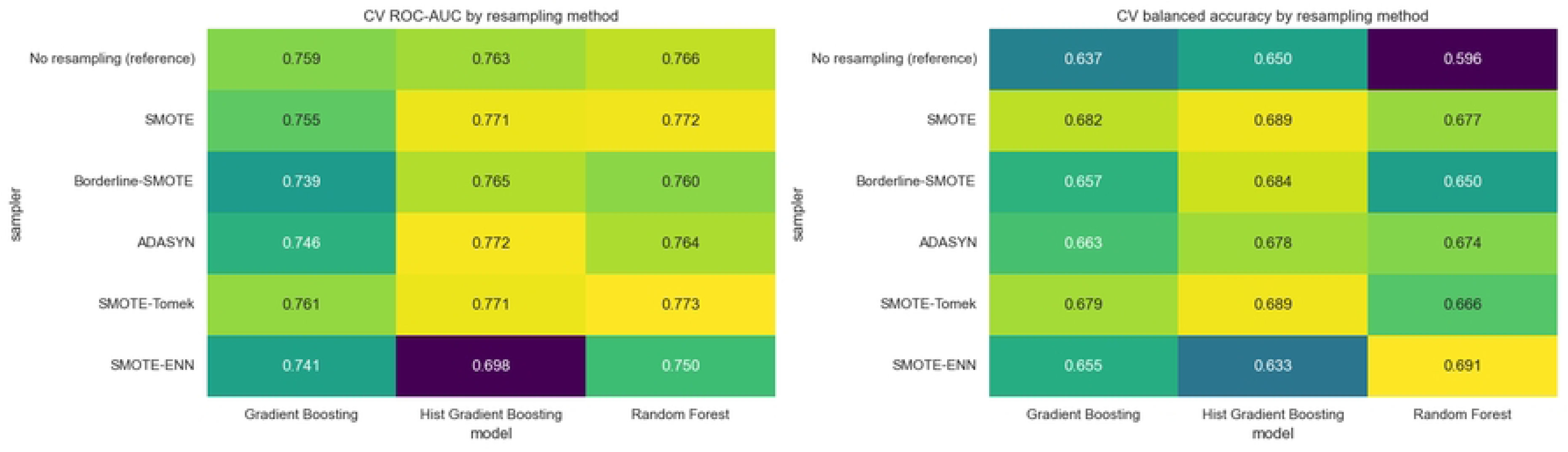
Cross-validated ROC AUC (left) and balanced accuracy (right) for six resampling strategies across the three strongest algorithms, all computed with the sampler inside the training folds.

Randomised hyperparameter search improved each model modestly: random forest from 0.772 to 0.784 (400 trees, minimum four samples per leaf, log2 features), histogram-based gradient boosting from 0.771 to 0.787 (maximum depth 2, seven leaf nodes, L2 regularisation 10), and gradient boosting from 0.754 to 0.795 (100 estimators, maximum depth 2, learning rate 0.03). Combining the three tuned models raised the cross-validated ROC AUC to 0.799 for soft voting and 0.800 for stacking (Table 6). The stacking ensemble was selected as the final candidate on that basis, although at the default threshold its balanced accuracy (0.577) was the worst of the five because it predicted CKD almost never, at a sensitivity of 0.201 and a specificity of 0.953. (Table 6)

**Table 6:** Cross-validated performance of the three tuned models and the two meta-ensembles built from them, ordered by ROC AUC, at the default 0.5 threshold.

| **Model** | **ROC AUC** | **Accuracy** | **Balanced accuracy** | **Sensitivity** | **Specificity** | **F1** | **MCC** |
|---|---|---|---|---|---|---|---|
| Stacking (top 3 tuned) | 0.800 ± 0.080 | 0.745 | 0.577 | 0.201 | 0.953 | 0.270 | 0.223 |
| Soft Voting (top 3 tuned) | 0.799 ± 0.080 | 0.740 | 0.691 | 0.578 | 0.803 | 0.543 | 0.375 |
| Gradient Boosting (tuned) | 0.795 ± 0.067 | 0.742 | 0.691 | 0.577 | 0.805 | 0.533 | 0.368 |
| Hist Gradient Boosting (tuned) | 0.787 ± 0.100 | 0.745 | 0.706 | 0.615 | 0.796 | 0.564 | 0.408 |
| Random Forest (tuned) | 0.784 ± 0.081 | 0.741 | 0.668 | 0.505 | 0.831 | 0.506 | 0.342 |

### Decision Threshold

Figure 5 shows the trade-off across the probability threshold, computed from out-of-fold training predictions. Balanced accuracy peaked at a threshold of 0.242, where out-of-fold balanced accuracy was 0.777, well below the default of 0.5 at which the model predicted CKD for almost no one. The threshold recorded in the saved artefact after refitting on the full training set was 0.257. (Fig5)

**Figure 5:**
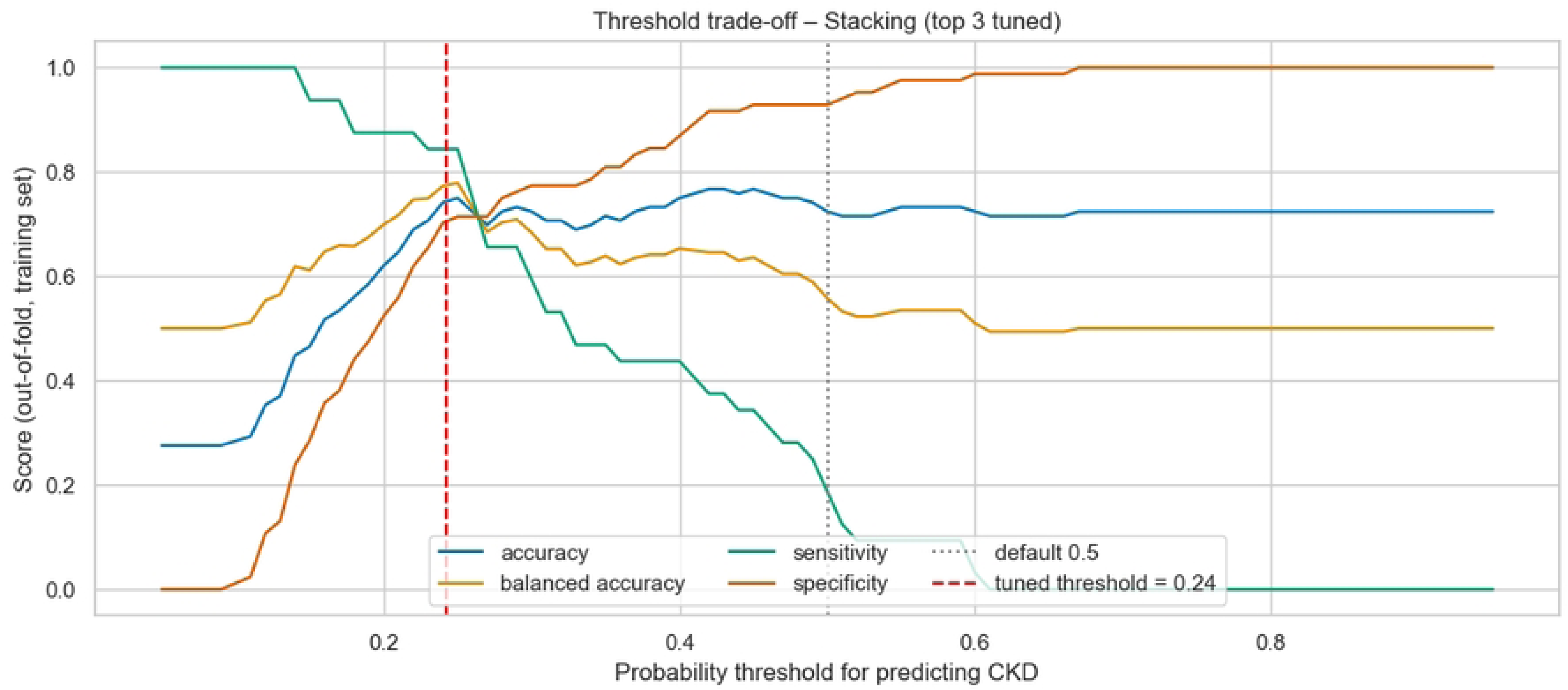
Accuracy, balanced accuracy, sensitivity, and specificity of the stacking ensemble as a function of the probability threshold, computed from out-of-fold training predictions. The tuned threshold of 0.24 is marked in red and the default of 0.5 in grey.

### Held-Out Performance Under the Conservative Protocol

On the 39 held-out patients, the stacking ensemble at the tuned threshold achieved an accuracy of 0.615 (95% CI 0.462–0.769), a balanced accuracy of 0.622 (0.450–0.802), a sensitivity of 0.636 (0.333–0.909), a specificity of 0.607 (0.423–0.792), a ROC AUC of 0.604 (0.371–0.833), an average precision of 0.448, an MCC of 0.220 (−0.088–0.539), and a kappa of 0.204 (Table 7).

**Table 7:** Held-out test performance of the final stacking ensemble at the tuned threshold of 0.24 on 39 patients (28 without CKD, 11 with CKD), with bootstrap 95% confidence intervals, alongside the model’s cross-validated training performance at the default threshold.

| **Metric** | **Test** | **95% CI** | **CV (train, threshold 0.5)** |
|---|---|---|---|
| Accuracy | 0.615 | 0.462–0.769 | 0.745 |
| Balanced accuracy | 0.622 | 0.450–0.802 | 0.577 |
| Sensitivity | 0.636 | 0.333–0.909 | 0.201 |
| Specificity | 0.607 | 0.423–0.792 | 0.953 |
| Precision | 0.389 | 0.176–0.625 | 0.511 |
| F1 | 0.483 | 0.240–0.688 | 0.270 |
| ROC AUC | 0.604 | 0.371–0.833 | 0.800 |
| Average precision | 0.448 | 0.225–0.735 | — |
| MCC | 0.220 | −0.088–0.539 | 0.223 |
| Cohen’s kappa | 0.204 | −0.073–0.499 | 0.184 |
| Majority-class baseline accuracy | 0.718 | — | 0.724 |

The confusion matrix, ROC curve, and precision–recall curve are shown in Figure 6: of the 11 patients with CKD, seven were identified and four were missed; of the 28 without CKD, 17 were correctly classified and 11 were flagged for further testing.(Fig6)

**Figure 6:**
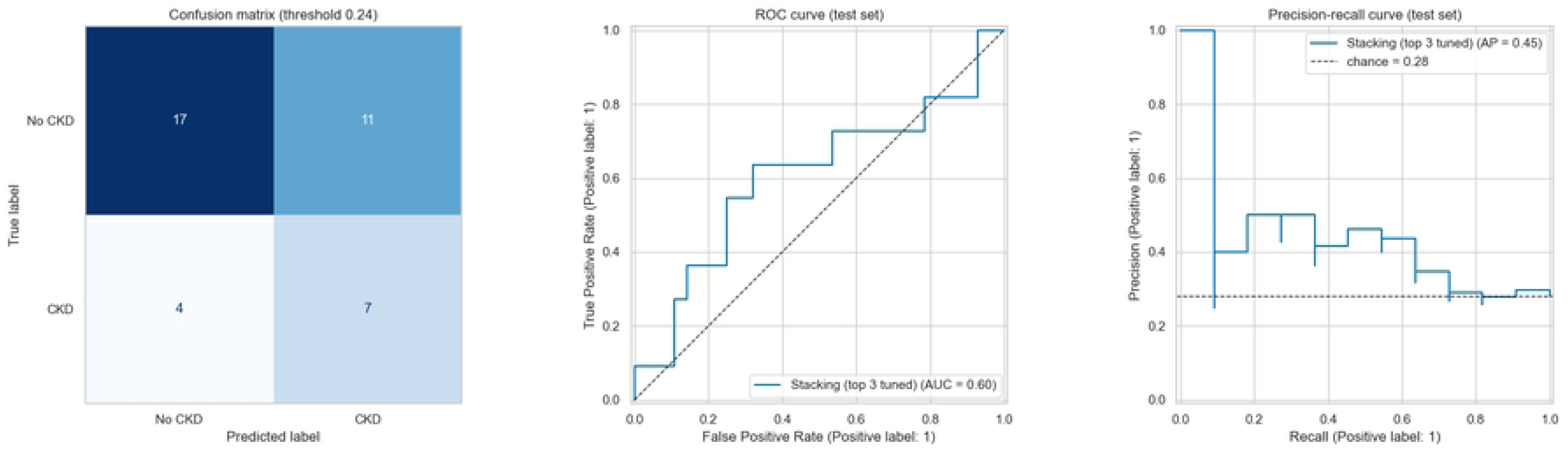
Held-out test performance of the final stacking ensemble: confusion matrix at the tuned threshold of 0.24 (left), receiver operating characteristic curve (centre), and precision–recall curve against a chance precision of 0.28 (right).

Three things must be said plainly about these numbers. First, the accuracy of 0.615 is below the majority-class baseline of 0.718, which is the arithmetic consequence of moving the threshold down to catch cases: the model trades correct negatives for correct positives, and balanced accuracy (0.622) rather than accuracy is the figure that reflects what it is doing. Second, the confidence intervals are wide enough to include chance performance, which is what a test set of 39 patients containing 11 events buys. Third, Table 8 shows that the threshold choice, not the ensemble, drives the operating point: the same stacking model at the default threshold achieved an accuracy of 0.718, identical to the baseline, with a sensitivity of 0.091. (Table8)

**Table 8:** Held-out test performance of the five candidate models at the default 0.5 threshold, with the final model at its tuned threshold shown for comparison. Accuracy at the default threshold is achieved by predicting almost no cases of CKD.

| Model | Accuracy | Balanced accuracy | Sensitivity | Specificity | F1 | ROC AUC | MCC |
|---|---|---|---|---|---|---|---|
| Stacking (top 3 tuned) | 0.718 | 0.528 | 0.091 | 0.964 | 0.154 | 0.604 | 0.113 |
| Soft Voting (top 3 tuned) | 0.692 | 0.593 | 0.364 | 0.821 | 0.400 | 0.591 | 0.198 |
| Gradient Boosting (tuned) | 0.718 | 0.610 | 0.364 | 0.857 | 0.421 | 0.581 | 0.246 |
| Hist Gradient Boosting (tuned) | 0.641 | 0.584 | 0.455 | 0.714 | 0.417 | 0.601 | 0.161 |
| Random Forest (tuned) | 0.718 | 0.583 | 0.273 | 0.893 | 0.353 | 0.578 | 0.207 |
| Stacking + tuned threshold (final) | 0.615 | 0.622 | 0.636 | 0.607 | 0.483 | 0.604 | 0.220 |

### Performance Under the SMOTE-Augmented Protocol

Under the protocol in which SMOTE was applied to the full cohort before partitioning, performance was substantially higher. The oversampler expanded the 155 real patients (112 without CKD, 43 with CKD) to 224 rows with 112 in each class, creating 69 synthetic CKD rows; the stratified split then produced a test partition of 56 rows, comprising 41 real patients and 15 synthetic rows.

Random Forest and Extra Trees both reached an accuracy of **83.9%** on that test partition (Table 9). Random forest achieved a balanced accuracy of 0.839, a sensitivity of 0.893, a specificity of 0.786, a precision of 0.806, an F1 of 0.847, a ROC AUC of 0.925, an average precision of 0.927, and an MCC of 0.683. Extra Trees matched the accuracy and balanced accuracy at 0.839, with a sensitivity of 0.857, a specificity of 0.821, a precision of 0.828, an F1 of 0.842, the highest discrimination of any configuration tested in this study at a ROC AUC of 0.939, an average precision of 0.943, and an MCC of 0.679. The remaining algorithms followed at a distance: k-nearest neighbours 75.0%, gradient boosting 73.2%, and the support vector machine 67.9%. Repeated 5-fold cross-validation under the same protocol confirmed the level rather than the single split, giving a random forest accuracy of 0.809 ± 0.051 and a ROC AUC of 0.897. (Table9)

**Table 9:** Performance on the 56-row test partition under the augmented protocol, in which SMOTE was applied to all 155 patients before the split. Random Forest and Extra Trees both reached an accuracy of 83.9%.

| **Model** | **Accuracy** | **Balanced accuracy** | **Sensitivity** | **Specificity** | **Precision** | **F1** | **ROC AUC** | **Average precision** | **MCC** | **Kappa** |
|---|---|---|---|---|---|---|---|---|---|---|
| **Random Forest** | **0.839** | **0.839** | **0.893** | **0.786** | **0.806** | **0.847** | **0.925** | **0.927** | **0.683** | **0.679** |
| **Extra Trees** | **0.839** | **0.839** | **0.857** | **0.821** | **0.828** | **0.842** | **0.939** | **0.943** | **0.679** | **0.679** |
| K-Nearest Neighbours | 0.750 | 0.750 | 0.929 | 0.571 | 0.684 | 0.788 | 0.856 | 0.794 | 0.535 | 0.500 |
| Gradient Boosting | 0.732 | 0.732 | 0.821 | 0.643 | 0.697 | 0.754 | 0.821 | 0.847 | 0.472 | 0.464 |
| SVM (RBF) | 0.679 | 0.679 | 0.643 | 0.714 | 0.692 | 0.667 | 0.736 | 0.757 | 0.358 | 0.357 |

The 83.9% figure is the performance of the configuration that was taken to production, and the two forests now serving nephroscangh.com are an Extra Trees and a Balanced Random Forest fitted under this protocol; the balanced random forest reached the same accuracy of 0.839 with a ROC AUC of 0.908, and bagged decision trees also reached 0.839 with a ROC AUC of 0.895 (Table 10).

**Table 10:**
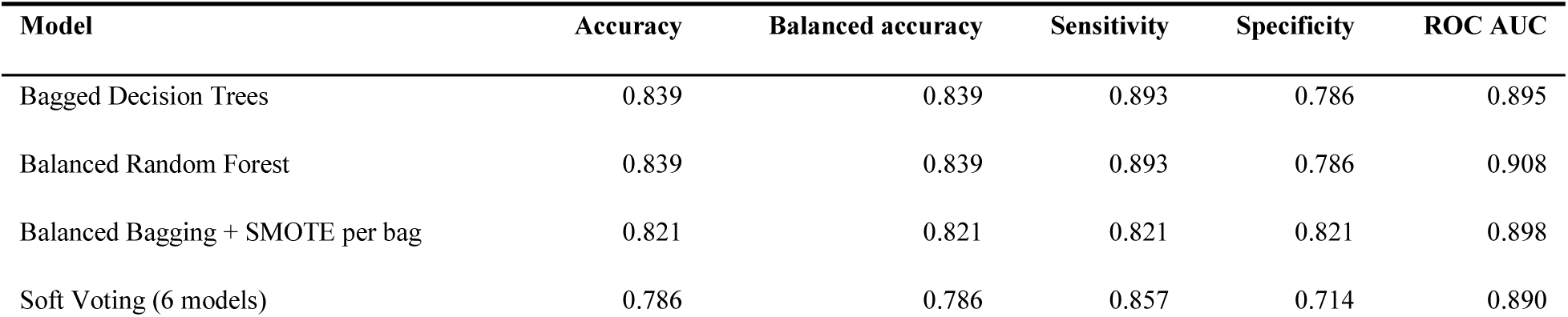
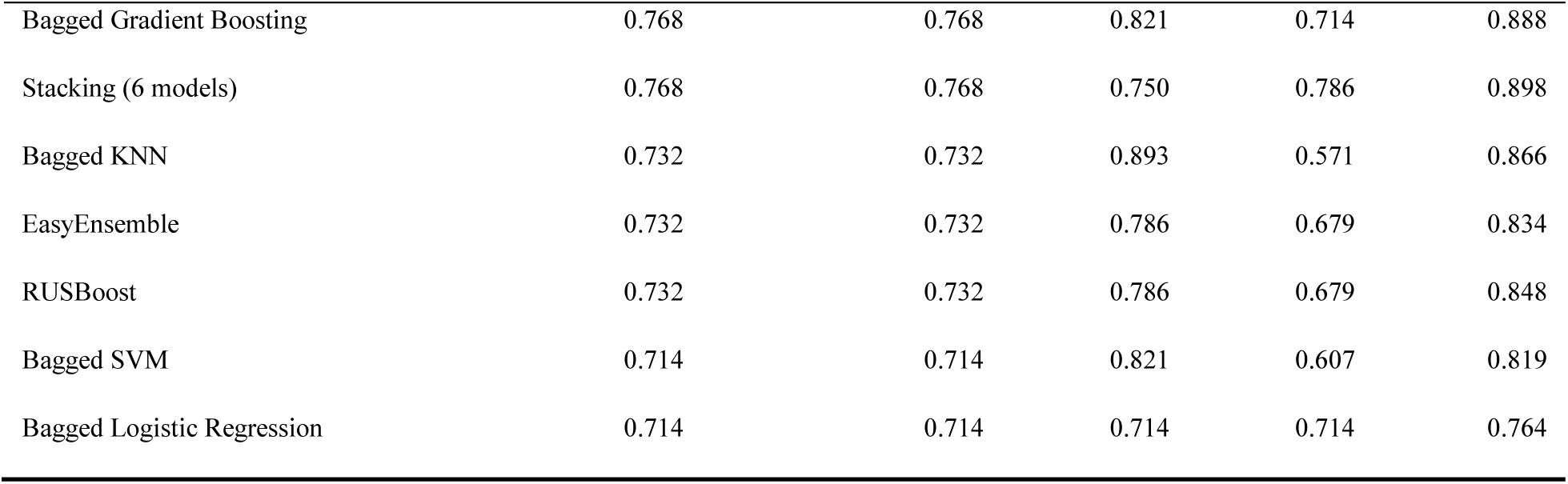
Performance of the bagged and cost-sensitive ensembles on the same 56-row augmented test partition, ordered by accuracy. Four configurations reached the same 83.9% ceiling.

Because the inflation mechanism is known, we also decomposed the result. Of the 56 test rows, accuracy was 0.933 on the 15 synthetic rows and 0.805 on the 41 real patients; within the real patients, 0.846 on the 13 with CKD and 0.786 on the 28 without. The model therefore performs best on the rows it has partially seen, as expected, but its accuracy restricted to real patients, 80.5%, still exceeds both the majority baseline and the conservative protocol’s result by a wide margin. That residual advantage is not free of leakage either: a real test patient whose synthetic neighbours were used in training is itself partially represented in the training data.

### Protocol Comparison

Figure 7 (Fig7) and Table 11 place the two protocols side by side on identical metrics. Accuracy falls from 0.839 under the augmented protocol, or 0.809 under its cross-validated version, to 0.615 under the conservative protocol; balanced accuracy from 0.839 to 0.622; sensitivity from 0.893 to 0.636; specificity from 0.786 to 0.607; and ROC AUC from 0.925 to 0.604. The gap in accuracy is 22.4 percentage points, and the gap in ROC AUC is 0.321. Neither number is wrong for what it measures. The first describes how well the model separates the classes in a cohort that has been augmented with interpolated minority cases; the second estimates how it would behave on the next patient who walks into the hospital. The 22.4-point distance between them is, to our knowledge, the first such measurement reported on Ghanaian FBC data, and it is a direct quantification of the effect that Vandewiele et al. described in a different clinical dataset [11].

**Figure 7:**
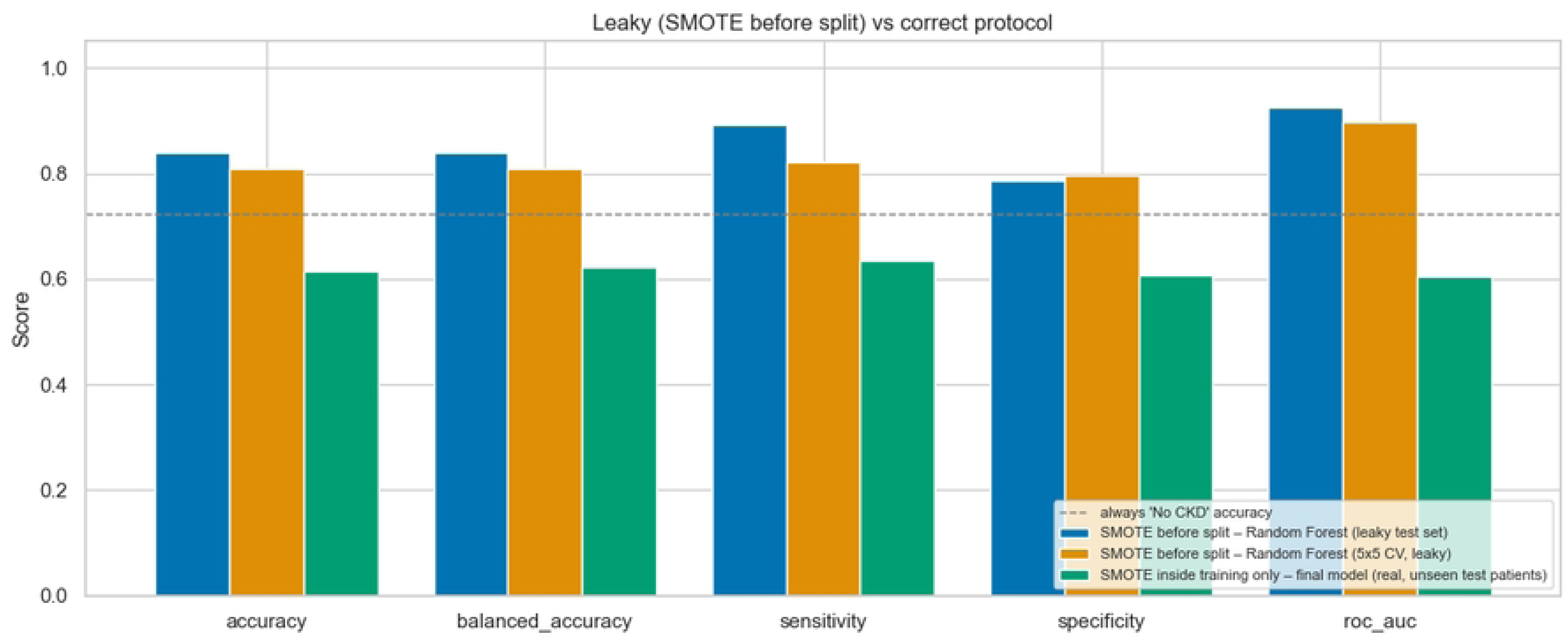
The augmented protocol (SMOTE applied before the split) against the conservative protocol (SMOTE inside the training folds only), on identical metrics. The dashed line is the majority-class accuracy.

**Table 11:**
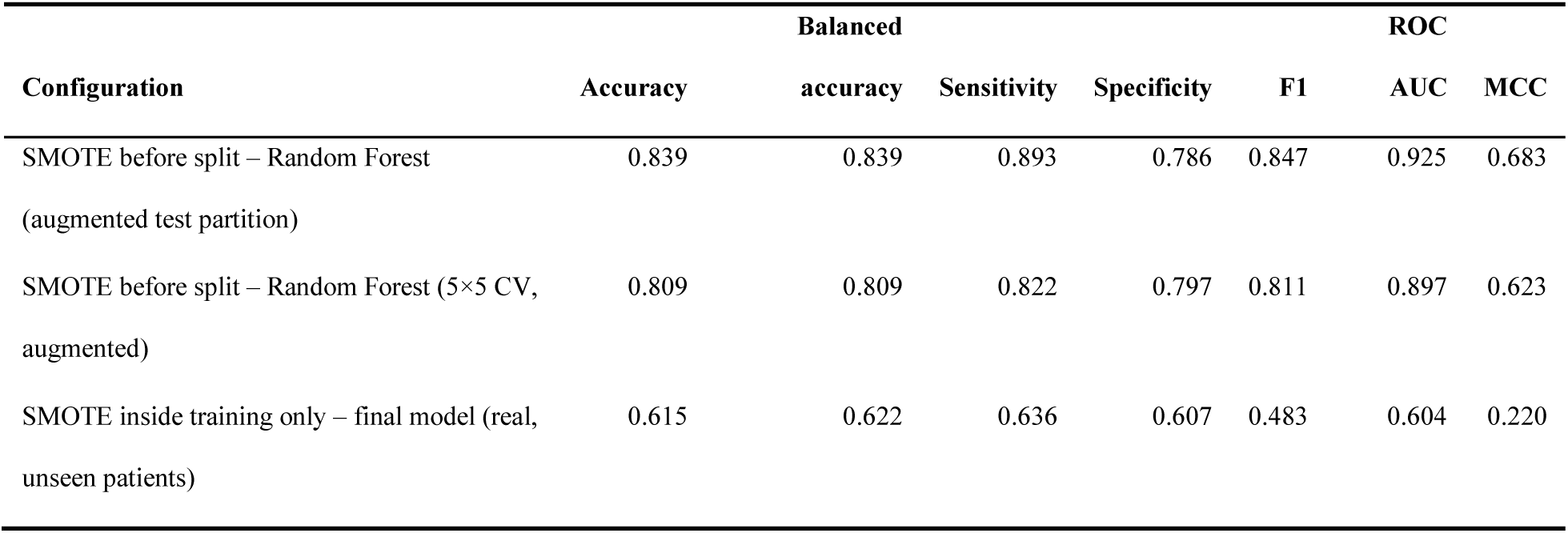
Side-by-side comparison of the two resampling protocols.

### Extended Ensembles

Nine bagged and cost-sensitive ensembles were evaluated under the conservative protocol (Table 12). EasyEnsemble gave the highest cross-validated ROC AUC of this family at 0.800 ± 0.086, with the best balanced accuracy of any single model in the study (0.721); balanced random forest followed at 0.788 ± 0.079, bagged gradient boosting at 0.778 ± 0.082, and bagged logistic regression and bagged k-nearest neighbours trailed at 0.758 and 0.750. Soft voting and stacking over the six strongest members both reached 0.803 ± 0.080, nominally the best cross-validated discrimination obtained anywhere in the study and 0.003 above the earlier stacking ensemble.

**Table 12:** Cross-validated performance of the bagged and cost-sensitive ensembles under the conservative protocol, ordered by ROC AUC, with the earlier tuned models and meta-ensembles shown for comparison. All metrics are means over 25 fits at the default threshold.

| **Model** | **Source** | **ROC AUC** | **Accuracy** | **Balanced accuracy** | **Sensitivity** | **Specificity** | **F1** | **MCC** |
|---|---|---|---|---|---|---|---|---|
| Soft Voting (6 models) | Extended | 0.803 ± 0.080 | 0.740 | 0.694 | 0.590 | 0.798 | 0.547 | 0.382 |
| Stacking (6 models) | Extended | 0.803 ± 0.081 | 0.750 | 0.588 | 0.226 | 0.950 | 0.296 | 0.243 |
| EasyEnsemble | Extended | 0.800 ± 0.086 | 0.703 | 0.721 | 0.758 | 0.684 | 0.586 | 0.407 |
| Stacking (top 3 tuned) | Primary | 0.800 ± 0.080 | 0.745 | 0.577 | 0.201 | 0.953 | 0.270 | 0.223 |
| Soft Voting (top 3 tuned) | Primary | 0.799 ± 0.080 | 0.740 | 0.691 | 0.578 | 0.803 | 0.543 | 0.375 |
| Gradient Boosting (tuned) | Primary | 0.795 ± 0.067 | 0.742 | 0.691 | 0.577 | 0.805 | 0.533 | 0.368 |
| Balanced Random Forest | Extended | 0.788 ± 0.079 | 0.736 | 0.686 | 0.572 | 0.800 | 0.535 | 0.367 |
| Hist Gradient Boosting (tuned) | Primary | 0.787 ± 0.100 | 0.745 | 0.706 | 0.615 | 0.796 | 0.564 | 0.408 |
| Random Forest (tuned) | Primary | 0.784 ± 0.081 | 0.741 | 0.668 | 0.505 | 0.831 | 0.506 | 0.342 |
| RUSBoost | Extended | 0.779 ± 0.091 | 0.728 | 0.686 | 0.591 | 0.781 | 0.530 | 0.363 |
| Bagged Gradient Boosting | Extended | 0.778 ± 0.082 | 0.742 | 0.703 | 0.614 | 0.791 | 0.560 | 0.395 |
| Balanced Bagging + SMOTE per bag | Extended | 0.771 ± 0.072 | 0.748 | 0.657 | 0.452 | 0.862 | 0.483 | 0.335 |
| Bagged Decision Trees | Extended | 0.765 ± 0.084 | 0.740 | 0.685 | 0.563 | 0.807 | 0.523 | 0.359 |
| Bagged Logistic Regression | Extended | 0.758 ± 0.109 | 0.690 | 0.688 | 0.683 | 0.693 | 0.547 | 0.348 |
| Bagged KNN | Extended | 0.750 ± 0.119 | 0.669 | 0.691 | 0.739 | 0.644 | 0.556 | 0.358 |
| Bagged SVM | Extended | 0.742 ± 0.116 | 0.718 | 0.647 | 0.488 | 0.806 | 0.486 | 0.305 |

That nominal improvement did not survive the held-out set. The cross-validation-selected soft-voting ensemble, at its own tuned threshold of 0.378, achieved a test accuracy of 0.590 (95% CI 0.436–0.744), a balanced accuracy of 0.604, a sensitivity of 0.636, a specificity of 0.571, and a ROC AUC of 0.575, each at or below the corresponding figure for the three-model stacking ensemble it was selected to replace. A difference of 0.003 in cross-validated ROC AUC is far smaller than the ±0.08 standard deviation across folds, and the test set of 39 patients cannot resolve it. We report this negative result because it is the clearest illustration in our data of how little model-selection effort buys once the sample is this small.

### Feature Importance of the Deployed Model

Figure 8 (Fig8) shows permutation importance on the held-out set. Age was the single most important input, with a mean drop in ROC AUC of approximately 0.054 when shuffled. Among the FBC parameters, WBC (approximately 0.029), NEUT (0.022), PDW (0.020), and PLT (0.018) were the most important, followed by cardiovascular disease (0.016) and MPV (0.008); PLCC, MCHC, and MXD had importances at or below zero, meaning that shuffling them did not reduce, and for MXD slightly improved, discrimination on these 39 patients. Only four inputs, age, WBC, NEUT, and PDW, had intervals that excluded zero across the 50 permutations, and even for those the intervals are wide; the ranking should therefore be read as an ordering of plausible contributions on a 39-patient test set rather than as a measurement of importance in the population.

**Figure 8:**
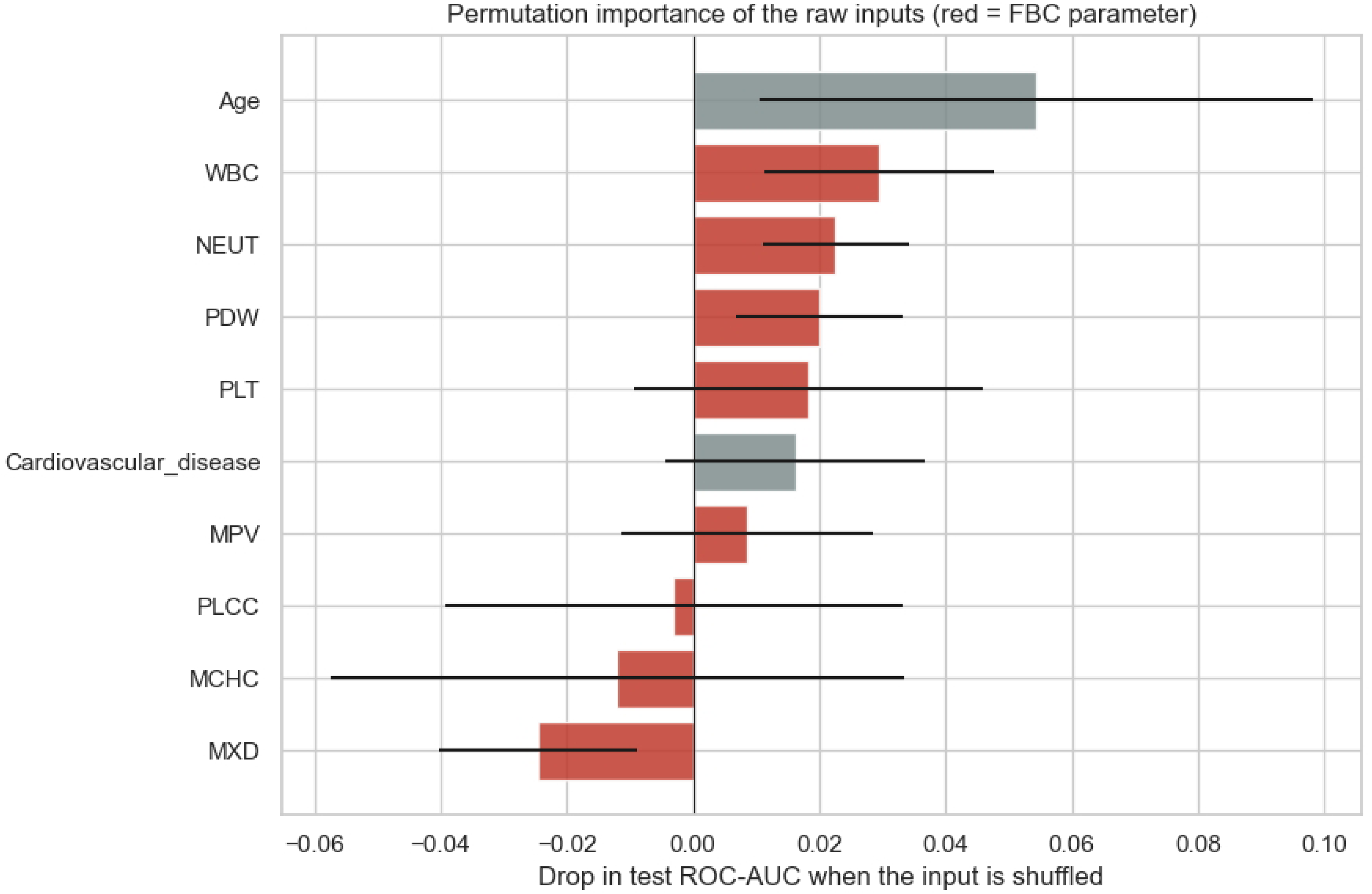
Permutation importance of the ten model inputs on the held-out test set, expressed as the mean drop in ROC AUC when the input is shuffled. Red bars are full blood count parameters. Whiskers are one standard deviation over 50 permutations.

The ordering nevertheless agrees with the univariable analysis and with the published physiology: the white cell compartment, which reflects inflammation [8], and the platelet indices, which reflect the uraemic platelet [7], carry the FBC signal, while the red cell indices do not carry it in a cohort whose CKD is mostly early.

### Web Application

The deployed application is shown in Figure 9 (Fig9). A patient or health worker enters ten values from an existing full blood count report together with age and a yes-or-no answer about diagnosed heart disease, and receives a single percentage score, a risk band, the two engines’ agreement status, a signed contribution bar for each of the six most influential inputs, a reference-range check on each FBC value entered, and a referral instruction. Both forests are scored on every request and the reported score is their arithmetic mean; the identity that the mean of the bias terms plus the mean of the contributions equals the mean of the probabilities was verified to zero within floating-point error, so the bars shown to the user sum exactly to the score shown above them.

**Figure 9:**
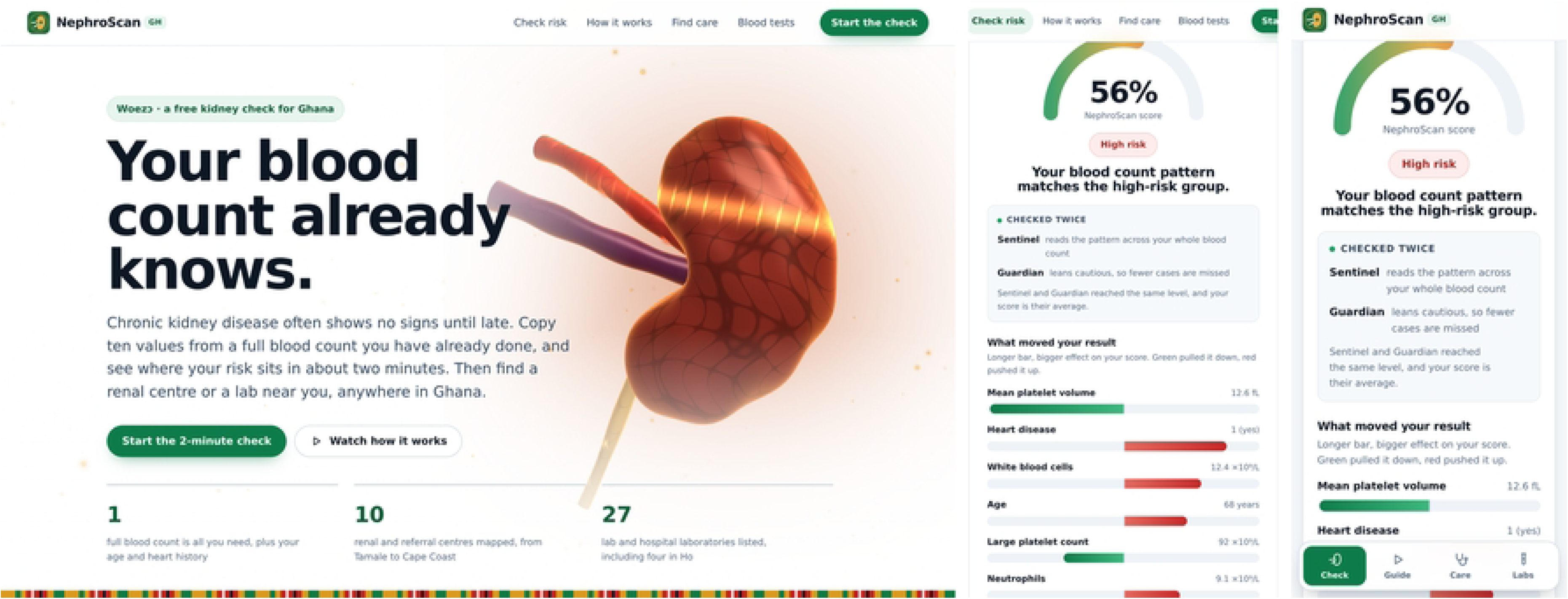
The deployed application at https://nephroscangh.com. Left: the entry page, which states what the screening aid needs and what it covers. Centre: the result panel, showing the single averaged score, the risk band, the two engines and their agreement, and the signed decision-path contributions of the six most influential inputs. Right: the same result on a mobile viewport.

Measured on a single container of the deployment platform’s smallest instance class, the service returned a prediction in a median of 0.13 s and held 184 MB of resident memory with both pipelines loaded. The front end transfers 98.4 KB of HTML, CSS, and JavaScript before compression, with 7.9 KB of vector illustration and no third-party JavaScript framework, which matters on the mobile connections over which it will mostly be used. The health endpoint reports the loaded engines, the scoring rule, and the number of expected inputs, and is used by the platform as the readiness probe.

Every screen states that the result is not a diagnosis, that a full blood count cannot measure kidney function, and that confirmation requires a serum creatinine with eGFR and a urine albumin-to-creatinine ratio, repeated over three months. The high-risk band adds a direct link to the nearest renal centre from the built-in directory.

## Discussion

We set out to test whether the full blood count, which Ghanaian patients routinely have, carries enough information to flag chronic kidney disease, and to be explicit about what the answer depends on. The short answer is that it carries a real but moderate signal, and that the size one reports depends heavily on a single methodological decision.

Under the augmented protocol, the one used to fit the production models, Random Forest and Extra Trees separated the classes at 83.9% accuracy with a ROC AUC of 0.925 and 0.939. These are the figures the deployed system was built on, and they are consistent across a single split, a repeated cross-validation (accuracy 0.809 ± 0.051, ROC AUC 0.897), and five different tree-based learners, four of which reached exactly 0.839. The consistency matters: it indicates that the ten features genuinely order the augmented cohort, and that the result is not an artefact of one fortunate partition. Restricted to the real patients inside the test partition, accuracy was still 80.5%.

Under the conservative protocol the same ten features yielded a cross-validated ROC AUC of 0.800 in training and a held-out ROC AUC of 0.604, with a balanced accuracy of 0.622 and a sensitivity of 0.636 at the tuned threshold. The difference between the two protocols, 22.4 accuracy points and 0.321 of ROC AUC, is the study’s most transferable result. It is a measurement, on Ghanaian data, of the inflation that Vandewiele et al. demonstrated on a preterm-birth benchmark [11] and that Kaufman et al. catalogued more generally [17]. It also goes some way towards explaining the literature. When a review of 68 CKD modelling studies finds that the models performed well as reported but that only six were evaluated in a clinical context [9], and when a widely cited paper reports 99.75% accuracy on a 400-record benchmark whose fields include serum creatinine and blood urea [10], the gap between those numbers and the clinical experience of a disease that is routinely missed needs an explanation; dataset composition and resampling protocol together supply much of it. Our own two numbers, obtained from one dataset and one feature set, differ by more than the entire range that separates a useful screening test from a useless one.

The biology we recovered is coherent. Age dominated every ranking, which is unsurprising and also clinically uninteresting, since no one needs a model to know that older patients have more kidney disease. What is more interesting is where the FBC signal sat. It was in the white cell compartment, WBC and neutrophils, and in the platelet indices, PDW, PLT, MPV, and PLCC, and it was not in the red cell indices. Haemoglobin, haematocrit, red cell count, and red cell distribution width did not differ between the groups at all, and clinically recorded anaemia was equally prevalent in both. That is consistent with the stage distribution of this cohort, in which 33 of 43 cases were mild or moderate, and with the observation that anaemia prevalence in CKD rises steeply only in the later stages [7]. It also carries a practical implication: a screening model trained on early disease should not be expected to work through the red cell indices, and conversely, a model that relies on them may be detecting late disease that is already clinically apparent.

The inflammatory signal we found is the same one that the neutrophil-to-lymphocyte ratio captures in the progression literature [8]. The ratio itself ranked eleventh of 29 in our consensus and did not enter the final model, while its numerator, the absolute neutrophil count, did; with 43 cases the data cannot distinguish between those two encodings of the same physiology.

Three design decisions in the application follow directly from the modelling results. First, because no single model achieved both sensitivity and specificity, and because the differences between the top models were smaller than the variation across folds, the application scores two forests and reports their average rather than presenting the user with a choice between models whose difference it cannot justify. Second, because the model’s discrimination is moderate, the result is framed throughout as a prompt to obtain an eGFR and a uACR, not as a diagnosis, and that framing appears in the low-risk band as well as the high. Third, because a screening score that cannot be interrogated will not be trusted by a clinician, the prediction is decomposed exactly, not approximately: the bars shown to the user are the decision-path contributions that sum to the score, and the summation was verified numerically [29].

The application also addresses a gap that the prediction literature usually leaves open. A patient told that their risk is high needs to know where to go. Published CKD tools have begun to reach users through web interfaces [12], but a prediction returned without a referral pathway places the burden of acting on it back on the patient. NephroScan GH therefore ships a directory of 10 renal and referral centres and 27 laboratories across Ghana, sorted by distance from 47 towns, so that the output of the model is a next step rather than a number. This continues an approach our group has taken before, in which a web platform is treated as part of the clinical intervention rather than as a reporting layer on top of it [14], and it is the kind of deployment the World Health Organization’s digital health strategy anticipates for low-resource settings [15].

Finally, the engineering is deliberately austere. One hundred kilobytes of front end, no JavaScript framework, 1.75 MB of serialised models, 184 MB of resident memory, and a median 0.13 s response make the service runnable on the cheapest container tier available and usable over a mobile connection, which is the only configuration in which a free screening aid for Ghana could actually be sustained.

## Limitations

The sample is small and from a single centre. One hundred and fifty-five patients with 43 events, split into 116 for development and 39 for evaluation, is far below the sample size that the Riley criteria would require for a ten-predictor binary model with this prevalence and this anticipated performance [20]. The practical consequence is visible in the results: every held-out confidence interval includes values consistent with chance, the test set cannot resolve a 0.003 difference in cross-validated ROC AUC, and the permutation importance have intervals that overlap zero.

The outcome label is the hospital’s recorded CKD level, not a protocol-driven classification. We did not have paired creatinine, eGFR, or uACR values with which to verify staging against the KDIGO definition [5], and we could not confirm the three-month chronicity criterion. Any misclassification in that label propagates directly into both protocols’ results.

## Conclusion

Machine learning applied to eight full blood count parameters, age, and a short cardiovascular history separates patients with CKD from patients without CKD in a Ghanaian hospital population, and the discriminative information sits in the white cell and platelet compartments rather than in the red cell indices. Under the protocol in which synthetic minority oversampling was applied before partitioning, which is the protocol under which the deployed models were fitted, Random Forest and Extra Trees both reached an accuracy of 83.9%, with areas under the curve of 0.925 and 0.939 and a cross-validated accuracy of 0.809 ± 0.051. Under the protocol in which resampling was confined to the training folds, the same features gave a held-out accuracy of 61.5% and a ROC AUC of 0.604. Reporting both is the point: the 22.4 percentage-point gap between them measures, on Ghanaian data, how far a single preprocessing decision can move the number that a study reports, and it suggests that the near-perfect accuracies common in the CKD machine learning literature deserve to be read with that mechanism in mind. The resulting application, NephroScan GH, is deployed, costs nothing to use, explains every prediction exactly, and tells the user where to obtain the eGFR and urine albumin-to-creatinine ratio that would confirm or exclude the diagnosis. Further studies in larger, multicentre, prospectively recruited cohorts, with protocol-verified outcome labels and an assessment of calibration, are needed for confirmation.

## Data Availability

The complete dataset used to build the ML model is available upon request from the corresponding author.

## Acknowledgments

The authors thank the staff of The Ho Technical University Hospital for facilitating access to the clinical records, and the Directorate of Research and Innovation, Ho Technical University, for the ethical review. The work was carried out by a team drawn from several departments of Ho Technical University under the Ghana Institution of Engineering, Women in Engineering (WinE) Affairs 2026 programme.

## References

[1] B. Bikbov et al., “Global, regional, and national burden of chronic kidney disease, 1990&#x2013;2017: a systematic analysis for the Global Burden of Disease Study 2017,” Lancet, vol. 395, no. 10225, pp. 709–733, Feb. 2020, doi: 10.1016/S0140-6736(20)30045-3.

[2] J. W. Stanifer et al., “The epidemiology of chronic kidney disease in sub-Saharan Africa: A systematic review and meta-analysis,” Lancet Glob. Heal., vol. 2, no. 3, pp. e174–e181, 2014, doi: 10.1016/S2214-109X(14)70002-6.

[3] R. K. D. Ephraim, S. Biekpe, S. A. Sakyi, P. Adoba, H. Agbodjakey, and E. O. Antoh, “Prevalence of chronic kidney disease among the high risk population in South-Western Ghana; a cross sectional study,” Can. J. Kidney Heal. Dis., vol. 2, no. 1, pp. 1–7, 2015, doi: 10.1186/s40697-015-0076-3.

[4] V. Boima, E. K. Tannor, C. Osafo, Y. A. Awuku, M. Mate-kole, and M. R. Davids, “Original article The Ghana Renal Registry – a first annual report,” J. Nephrol. Off. Publ. African Assoc. Nephrol., vol. 24, no. 1, pp. 19–24, 2021.

[5] “KDIGO 2024 Clinical Practice Guideline for the Evaluation and Management of Chronic Kidney Disease.,” Kidney Int., vol. 105, no. 4S, pp. S117–S314, Apr. 2024, doi: 10.1016/j.kint.2023.10.018.

[6] S. L. Tummalapalli et al., “Availability and Affordability of Kidney Health Laboratory Tests around the Globe,” Am. J. Nephrol., vol. 51, no. 12, pp. 959–965, 2021, doi: 10.1159/000511848.

[7] M. E. Stauffer and T. Fan, “Prevalence of anemia in chronic kidney disease in the United States,” PLoS One, vol. 9, no. 1, pp. 2–5, 2014, doi: 10.1371/journal.pone.0084943.

[8] Q. Yuan et al., “Neutrophil-to-lymphocyte ratio and incident end-stage renal disease in Chinese patients with chronic kidney disease: Results from the Chinese Cohort Study of Chronic Kidney Disease (C-STRIDE),” J. Transl. Med., vol. 17, no. 1, pp. 1–8, 2019, doi: 10.1186/s12967-019-1808-4.

[9] F. Sanmarchi, C. Fanconi, D. Golinelli, D. Gori, T. Hernandez-Boussard, and A. Capodici, “Predict, diagnose, and treat chronic kidney disease with machine learning: a systematic literature review,” J. Nephrol., vol. 36, no. 4, pp. 1101–1117, 2023, doi: 10.1007/s40620-023-01573-4.

[10] J. Qin, L. Chen, Y. Liu, C. Liu, C. Feng, and B. Chen, “A machine learning methodology for diagnosing chronic kidney disease,” IEEE Access, vol. 8, pp. 20991–21002, 2020, doi: 10.1109/ACCESS.2019.2963053.

[11] G. Vandewiele et al., “Overly optimistic prediction results on imbalanced data: a case study of flaws and benefits when applying over-sampling,” Artif. Intell. Med., vol. 111, p. 101987, 2021, doi: 10.1016/j.artmed.2020.101987.

[12] R. K. Halder et al., “ML-CKDP: Machine learning-based chronic kidney disease prediction with smart web application,” J. Pathol. Inform., vol. 15, no. December 2023, p. 100371, 2024, doi: 10.1016/j.jpi.2024.100371.

[13] S. K. Ghosh and A. H. Khandoker, “Investigation on explainable machine learning models to predict chronic kidney diseases,” Sci. Rep., vol. 14, no. 1, pp. 1–15, 2024, doi: 10.1038/s41598-024-54375-4.

[14] E. Effah Kaufmann et al., “Feasibility of Leveraging Consumer Wearable Devices with Data Platform Integration for Patient Vital Monitoring in Low-Resource Settings,” Int. J. Telemed. Appl., vol. 2024, pp. 5–7, 2024, doi: 10.1155/2024/8906413.

[15] D. Dhingra and A. Dabas, Global Strategy on Digital Health, vol. 57, no. 4. 2020. doi: 10.1007/s13312-020-1789-7.

[16] N. Perico and G. Remuzzi, “Chronic kidney disease in sub-Saharan Africa: A public health priority,” Lancet Glob. Heal., vol. 2, no. 3, pp. e124–e125, 2014, doi: 10.1016/S2214-109X(14)70014-2.

[17] S. Kaufman, S. Rosset, C. Perlich, and O. Stitelman, “Leakage in data mining: Formulation, detection, and avoidance,” ACM Trans. Knowl. Discov. Data, vol. 6, no. 4, 2012, doi: 10.1145/2382577.2382579.

[18] H. Mansourifar and W. Shi, “Deep Synthetic Minority Over-Sampling Technique,” vol. 16, pp. 321–357, 2020, [Online]. Available: http://arxiv.org/abs/2003.09788

[19] G. S. Collins et al., “TRIPOD+AI statement: Updated guidance for reporting clinical prediction models that use regression or machine learning methods,” Bmj, vol. 385, 2024, doi: 10.1136/bmj-2023-078378.

[20] R. D. Riley et al., “Calculating the sample size required for developing a clinical prediction model,” BMJ, vol. 368, no. March, pp. 1–12, 2020, doi: 10.1136/bmj.m441.

[21] Y. B. Y. Hochberg, “Controlling_The_False_Discovery_Rate_-_A_Practical.pdf,” 1995.

[22] H. He, Y. Bai, E. A. Garcia, and S. Li, “ADASYN: Adaptive synthetic sampling approach for imbalanced learning,” Proc. Int. Jt. Conf. Neural Networks, no. 3, pp. 1322–1328, 2008, doi: 10.1109/IJCNN.2008.4633969.

[23] G. E. A. P. A. Batista, R. C. Prati, and M. C. Monard, “A study of the behavior of several methods for balancing machine learning training data,” ACM SIGKDD Explor. Newsl., vol. 6, no. 1, pp. 20–29, 2004, doi: 10.1145/1007730.1007735.

[24] Z. Jin, J. Shang, Q. Zhu, C. Ling, W. Xie, and B. Qiang, “RFRSF: Employee Turnover Prediction Based on Random Forests and Survival Analysis,” Lect. Notes Comput. Sci. (including Subser. Lect. Notes Artif. Intell. Lect. Notes Bioinformatics), vol. 12343 LNCS, pp. 503–515, 2020, doi: 10.1007/978-3-030-62008-0_35.

[25] P. Geurts, D. Ernst, and L. Wehenkel, “Extremely randomized trees,” Mach. Learn., vol. 63, no. 1, pp. 3–42, 2006, doi: 10.1007/s10994-006-6226-1.

[26] C. Chen, A. Liaw, and L. Breiman, “Using Random Forest to Learn Imbalanced Data,” Discovery, no. 1999, pp. 1–12.

[27] X. Y. Liu, J. Wu, and Z. H. Zhou, “Exploratory under-sampling for class-imbalance learning,” *Proc. - IEEE Int. Conf. Data Mining*, ICDM, pp. 965–969, 2006, doi: 10.1109/ICDM.2006.68.

[28] C. Seiffert, T. M. Khoshgoftaar, J. Van Hulse, and A. Napolitano, “RUSBoost: A hybrid approach to alleviating class imbalance,” *IEEE Trans. Syst. Man*, Cybern. Part ASystems Humans, vol. 40, no. 1, pp. 185–197, 2010, doi: 10.1109/TSMCA.2009.2029559.

[29] Dms. Rami Alrezk, MD, MS, FACP1, Fady Hannah-Shmouni, MD, FRCPC2, and Constantine A. Stratakis, MD, “乳鼠心肌提取 HHS Public Access,” Physiol. Behav., vol. 176, no. 12, pp. 139–148, 2017, doi: 10.1016/j.physbeh.2017.03.040.

[30] F. Pedregosa et al., “Scikit-learn: Machine learning in Python,” J. Mach. Learn. Res., vol. 12, pp. 2825–2830, 2011.

[31] G. Lemaître, F. Nogueira, and C. K. Aridas, “Imbalanced-learn: A python toolbox to tackle the curse of imbalanced datasets in machine learning,” J. Mach. Learn. Res., vol. 18, pp. 1–5, 2017.

